# Covid-19 Pandemic Data Analysis and Forecasting using Machine Learning Algorithms

**DOI:** 10.1101/2020.06.25.20140004

**Authors:** Sohini Sengupta, Sareeta Mugde, Garima Sharma

## Abstract

India reported its first Covid-19 case on 30th Jan 2020 and the number of cases reported heavily escalated from March, 2020. This research paper analyses COVID -19 data initially at a global level and then drills down to the scenario obtained in India. Data is gathered from multiple data sources-several authentic government websites. The need of the hour is to accurately forecast when the numbers will reach at its peak and then diminish. It will be of huge help to public welfare professionals to plan the preventive measures to be taken keeping the economic balance of the country as well. Variables such as gender, geographical location, age etc. have been represented using Python and Data Visualization techniques. Time Series Forecasting techniques including Machine Learning models like Linear Regression, Support Vector Regression, Polynomial Regression and Deep Learning Forecasting Model like LSTM(Long short-term memory) are deployed to study the probable hike in cases and in the near future. A comparative analysis is also done to understand which model fits the best for our data. Data is considered till 30^th^ July, 2020. The results show that a statistical model named sigmoid model is outperforming other models. Also the Sigmoid model is giving an estimate of the day on which we can expect the number of active cases to reach its peak and also when the curve will start to flatten. Strength of Sigmoid model lies in providing a count of date that no other model offers and thus it is the best model to predict Covid cases counts –this is unique feature of analysis in this paper. Certain feature engineering techniques have been used to transfer data into logarithmic scale as is affords better comparison removing any data extremities or outliers. Based on the predictions of the short-term interval, our model can be tuned to forecast long time intervals.

## Introduction

The Covid-19 pandemic in the past four months has engulfed India with huge surge in the number of patients afflicted with the virus as also increase in the number of fatalities attributed to the disease.

Unfortunately India currently reports the highest number of cases in Asia. However the fatality rate is relative lower (2.8%) as compared to the world’s rate of 6.1% (Source-Economic Times) as of 3rd June 2020. On 12th January 2020, the World Health Organization (WHO) declared the novel coronavirus was responsible for acute respiratory illness of a community of people in Wuhan, China. First confirmed case in India was reported on 30th, 2020 and the first confirmed death was reported on 12th March, 2020. Eventually all Indians were implored by the Government of India to maintain social distancing as a preventive measure. Nationwide Lockdown initiated a number of measures to contain the spread of COVID-19, prominent among them being-Only essential services were allowed to be open; Major cities and some states made wearing face-masks compulsory; Central armed forces came into action in a few areas;. Helpline numbers were set up to help patients or those showing signs of affliction. Various research agencies along with Government of India took the initiative to gather data on Covid-19 and a database was setup that included real time data of number of confirmed cases, deaths, recovery dates, break-up of cases basis age, gender, geographical location, as well as comparison of India’s position with respect to other countries. Further, data was also collected that included information on the number of diagnostic tests that were being conducted at a state and district level. The nation was divided into three zones namely a) Red zones (Hotspots) b) Orange zones (non-hotspots) and c) Green zone (districts without confirm cases for three consecutive weeks). The nation witnessed huge economic downfall in these months. Thousands of people lost jobs. Retail sector became the biggest casualty of lockdown. Also the tourism, hospitality and aviation sector faced massive losses owing to pandemic related regulations allowing near to zero inflows of tourists and visitors severely impacting top-lines of travel agents, hotel and aviation. Global supply chain has been immensely disturbed. Centre for Monitoring the Indian economy (CMIE) reported unemployment rate touching 30% in urban and 21% in rural areas bringing the total unemployment rate of the country to 23.8%.

## Review of Related Literature

Ever since the emergence of COVID -19 and it’s consequent spread across continents engulfing both advanced and developing nations, there has been a lot of research papers publications on various aspects of COVID-19. Apart from researches being undertaken in the domain of vaccination, drug therapy and other clinical aspects, considerable research work is also being carried out with patients as the fulcrum-patients who have recovered; patients with co-morbidities and the incidence of virus etc. Thorough analysis is being performed on the people who recovered so as to shed some light on how to deal with the active cases. Data scientists all over the world are busy in making sense out of the available data and predict the near future. Finding trend pattern, feature selection, forecasting techniques are being applied in and out to come to a conclusion. ^[1]^ Gupta, Pal and Kumar (2020) in their research paper ‘Trend analysis and forecasting of covid-19 break in India’ used exploratory data analysis to report the situation in the time period of January to March in India. They use time series forecasting methods to predict the future trends. A very famous machine learning model-Arima model prediction was used and inferred that a huge surge in the number of likely covid-19 positive cases was predicted in April and May. The average that was forecasted was a detection of approximately 7000 patients in a total span of 30 days in April. However in reality the figures were higher. ^[2]^Another research paper named SEIR and Regression Model based COVID-19 outbreak predictions in India’ by Pandey and Chowdhury (2020) from department of CSE and IT of Northcap University, India in collaboration with Defence Research and Development Organization (DRDO) India also covered data from January 30 to March 30, 2020. They used regression models for forecasting. According to them, expected cases may rise to about 5000 in a two week time period. This was far more accurate than the model predicted by Pal and Gupta however actual scenario showed a bigger upsurge. The findings of the paper may be relevant to several other sectors or other branches of healthcare as immunity power is strongly related to fighting with Covid-19. Healthcare experts also maintain that people with a less developed immunity system are more likely to be a victim of Covid-19. ^[3]^Dovey, Hurell, May (2005) in their Research Paper titled “Young adults’ (16-25 years) suggestions for providing developmentally appropriate diabetes services: A qualitative study. “ published in the journal of ‘Health Social Care in the community’ with focus on the opinion of young adults (16-25yrs) proffered suggestions for providing developmentally appropriate diabetic services and it is widely recognized that those suffering from diabetes are more prone to many other diseases owing to compromised immunity system. Hence it’s recommended that we gather information from healthcare workers on staying fit and healthy during this pandemic. ^[4]^Another very relevant paper named “Analysis of Spatial Spread Relationships of Coronavirus (COVID-19) Pandemic in the World using Self Organizing Maps (2020)“ by Julio, Monica, Sanchez, Castillo in the journal of ‘Chaos, Solitons & Fractals’ uses clustering methods to analyze countries on the basis of most affected patients and how they are reacting to it. This paper shows the world scenario first and thereby drilling down to the country of Mexico. Speaking of a country like India, where 65.97% (Trading Economics) belong to rural population, accessibility and utilization of primary health care during the current pandemic situation is a big question. ^[5]^ A similar study has been done in UK by Field, Briggs(2020) and we can find its related paper named “Socio-economic and locational determinants of accessibility and utilization of primary health-care” in the journal-‘Health Social Care in the community’. This inspires me to conduct a similar study in India. ^[6]^Another recently published paper by Singhal, Singh, Lall – “Modeling and prediction of COVID-19 pandemic using Gaussian mixture model, 2020” in the journal - ‘Chaos Solitons & Fractals’ captures the trend of cases and also a prediction using Fourier Decomposition Method. This paper also collects data till 30^th^ July and predicts the expected number of cases and deaths in the upcoming days.

## Analysis

Visualizations had always been easy to understand the raw data. Here we are going to compare the growth of Covid-19 confirmed, death and recovered cases of India to other major countries that have also been heavily infected. The visualizations are created in Python using the matplotlib, seaborn, plotly libraries and also datetime library for time series data analysis. Date of India and world is gathered from multiple data sheets from kaggle.

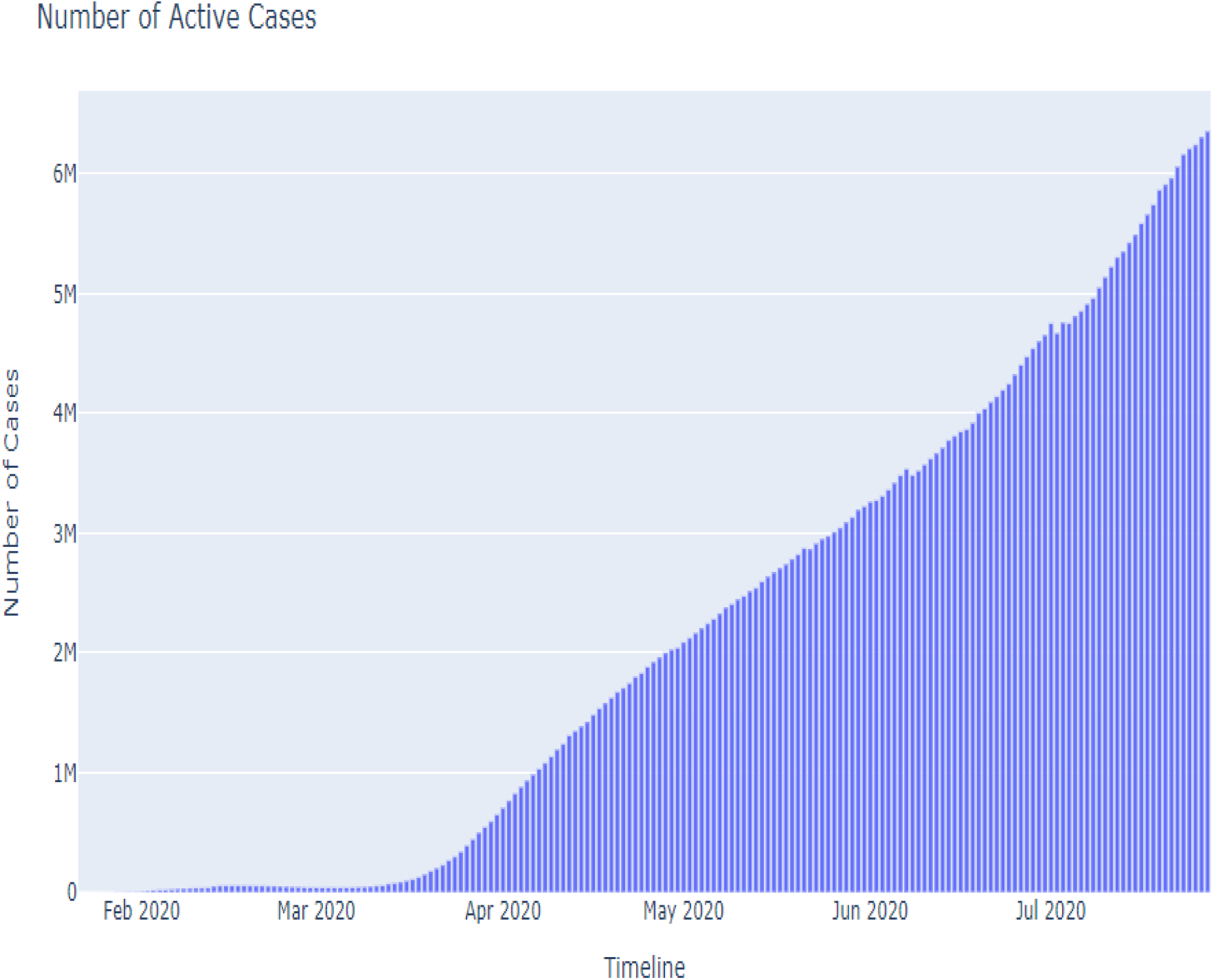

Active cases= confirmed cases-recovered cases-death cases

An increase in number of active cases signifies a considerable drop in the number of recovered and death case with respect to the number of confirmed cases. To further confirm this we can visualize the number of closed cases as well.

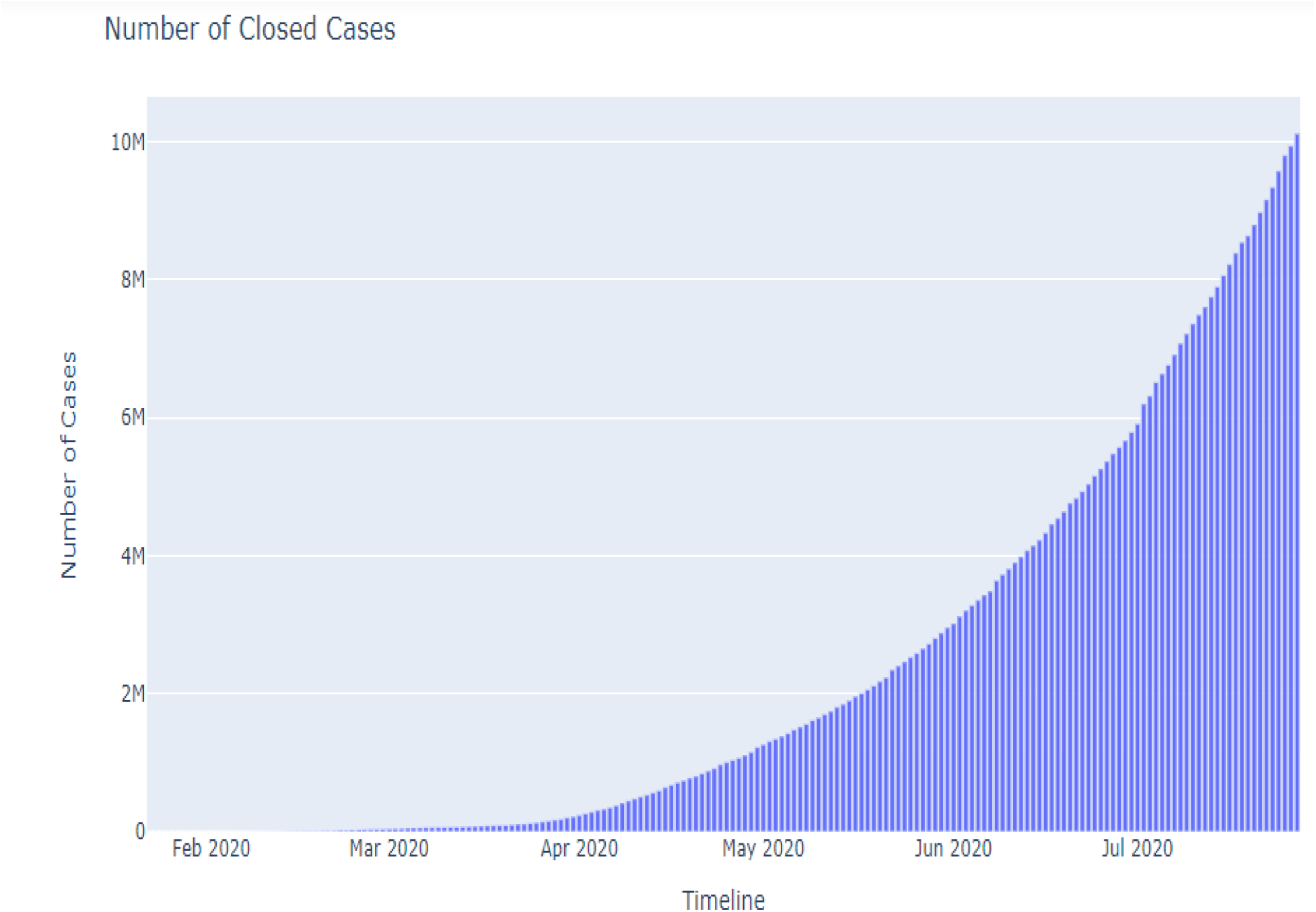

The above graph supports the fact that either more patients are getting recovered from the disease or more patients are dying due to Covid-19.

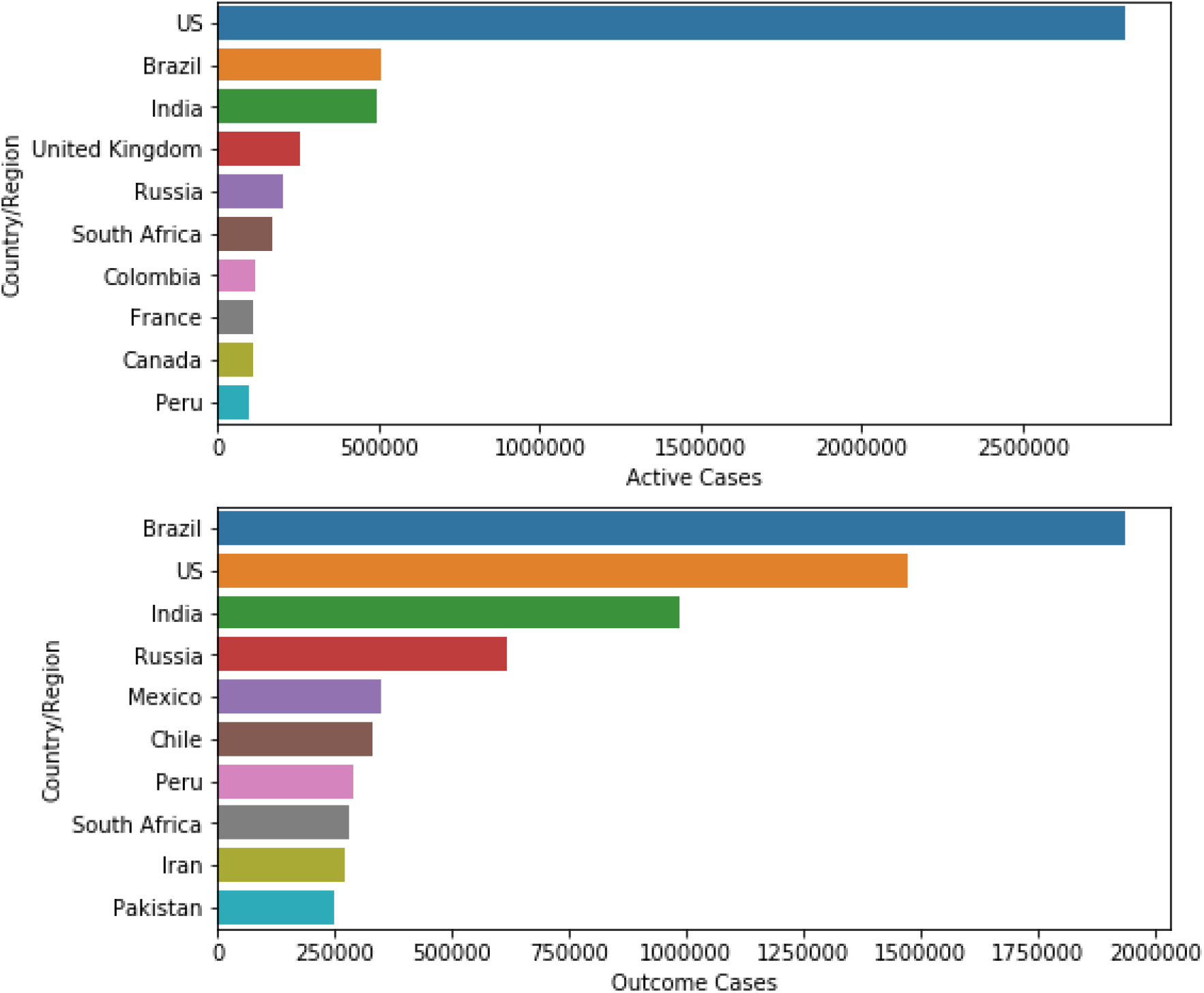

Here’s a bar graph to compare the active and closed cases of different countries as of 30^th^ July 2020. India’s active cases remain at 3^r^ position in the world it comes to the 3^r^ position in the number of closed cases.

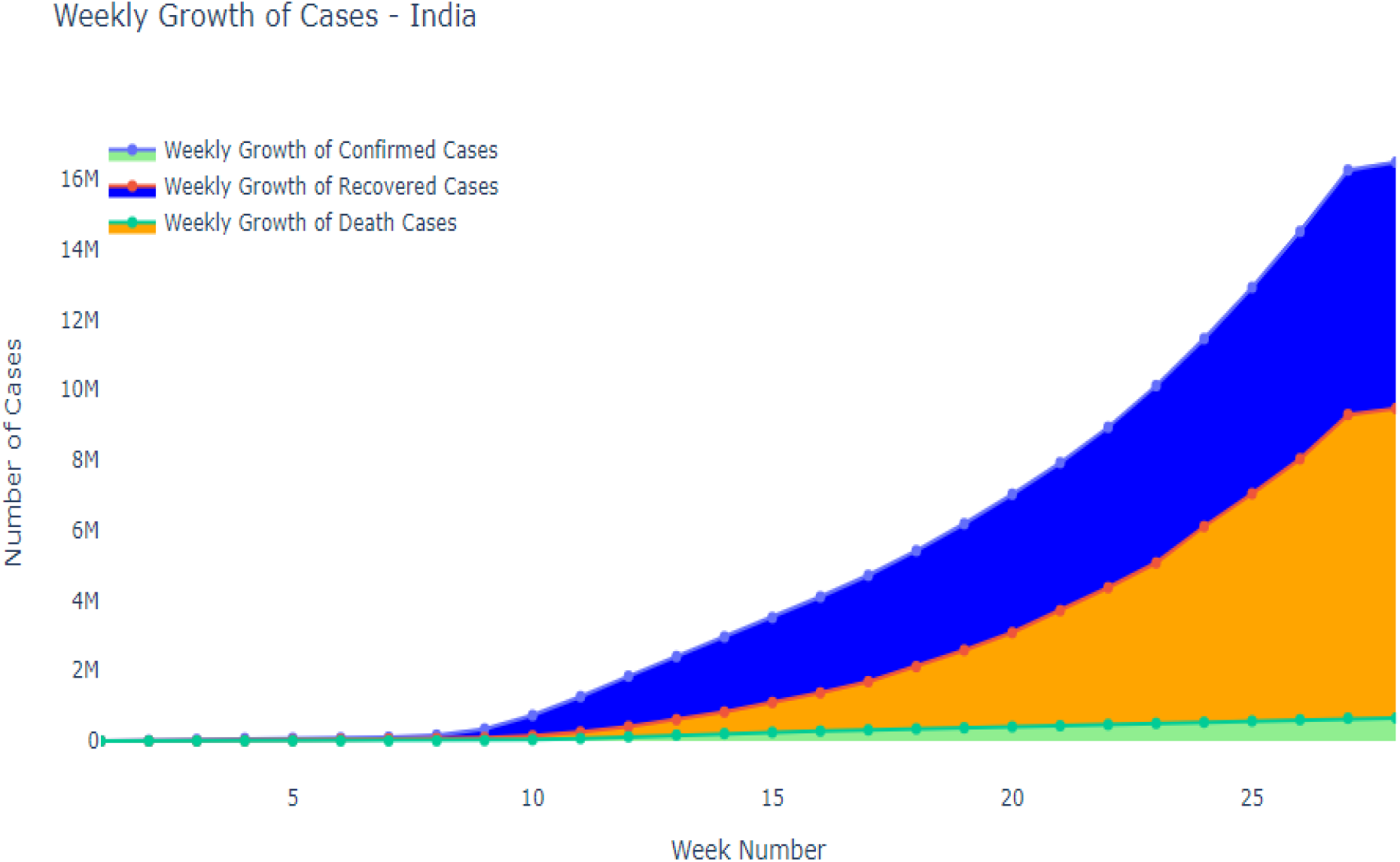

Weekly growth of cases worldwide starting from January 2020 cases considerable increase from week 9 that is March and experience sharp growth.

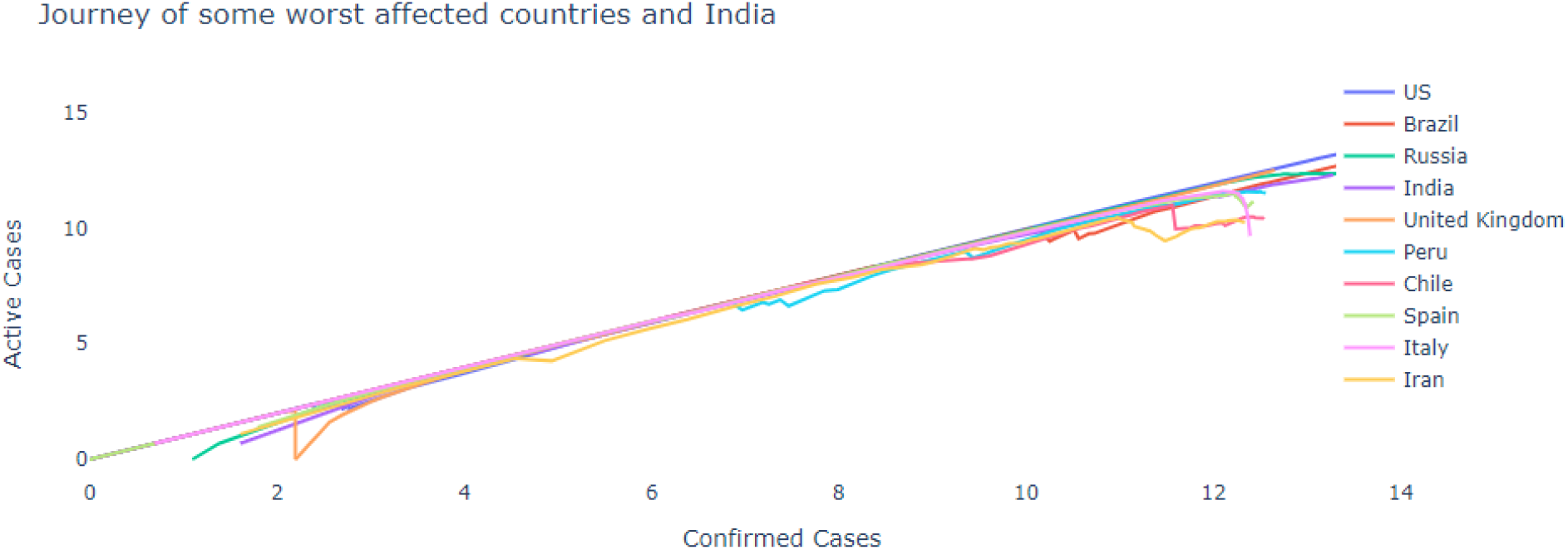

The visualization is done on a logarithmic scale. It is quite evident that the pandemic has spread in all the countries in same manner, however certain countries are practicing controlling procedures rigorously and it sevident from the graph. Most of the countries are following the same trajectories as US i.e uncontrolled exponential growth while a few countries like Iran, Spain, Italy, Chile have started showing a dip indicating signs of control over Covid-19.

Clustering –

Machine learning provides an excellent feature of clustering which will help us to categorize countries on the basis of severity of the pandemic. Severity can be measured on several features; here we are considering the mortality and recovery rate of countries. We are using k-means clustering and hierarchical clustering methods both of which suggests that suitable number of clusters will be 3.

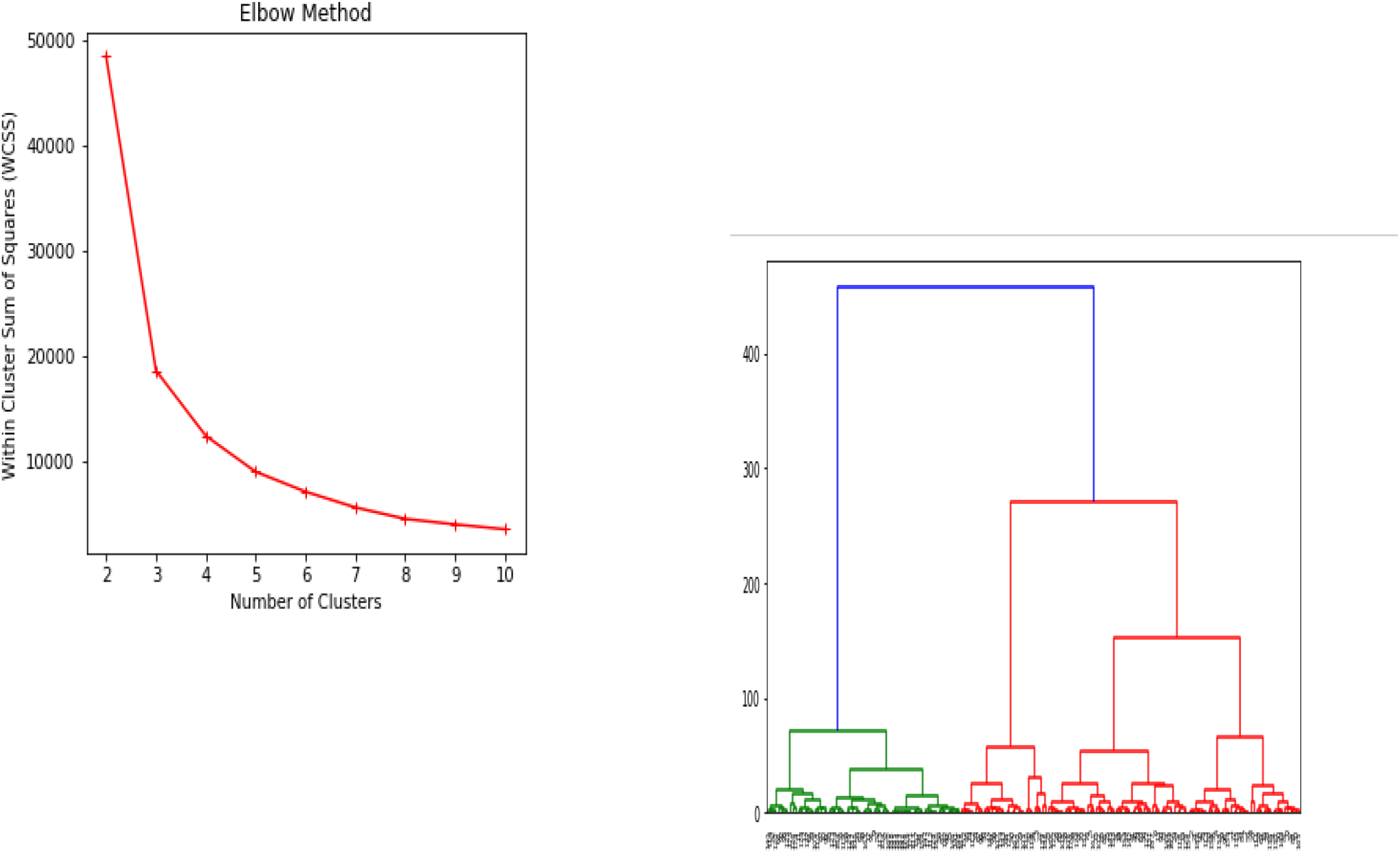
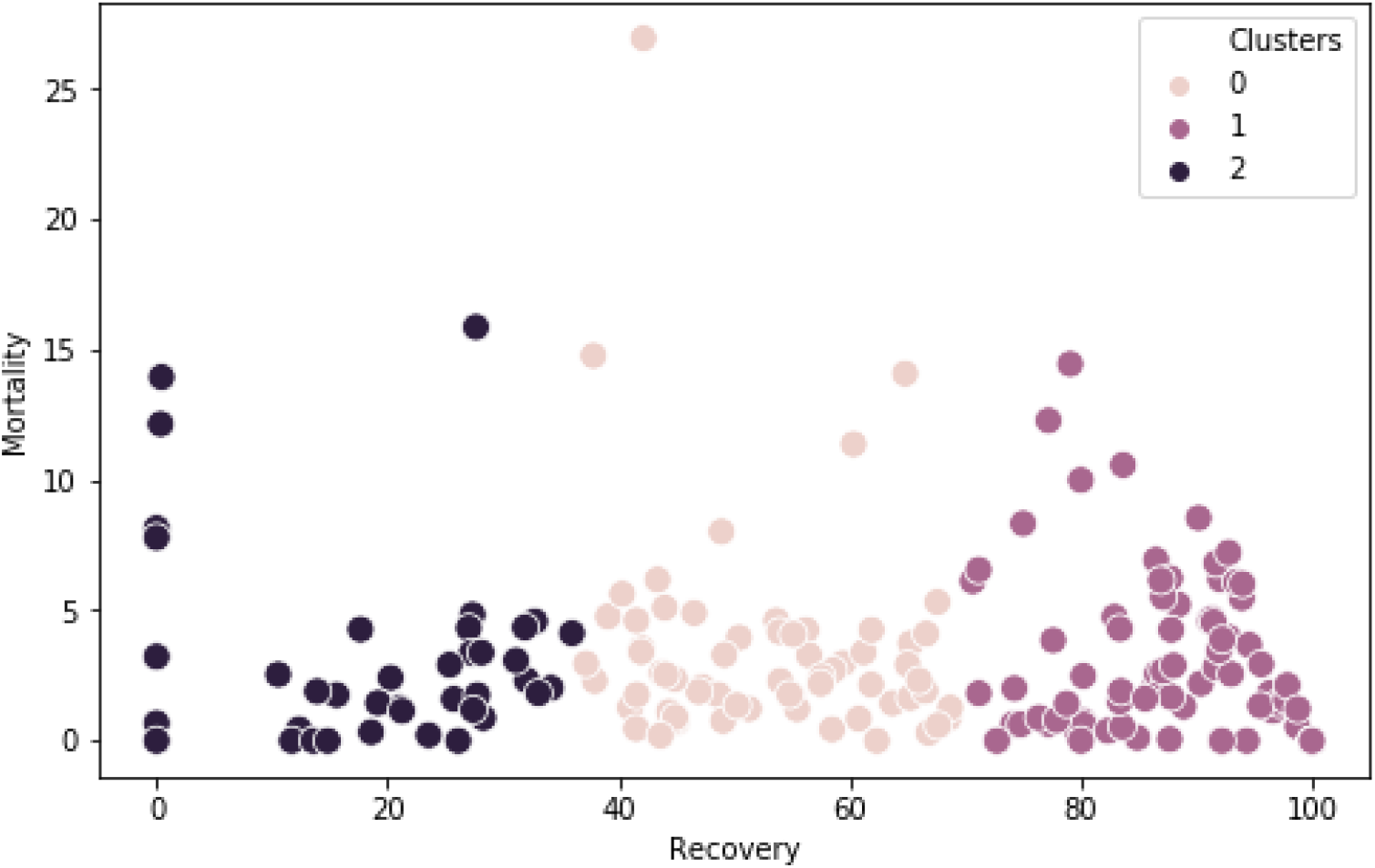

Therefore considering k= 3, we get the following.

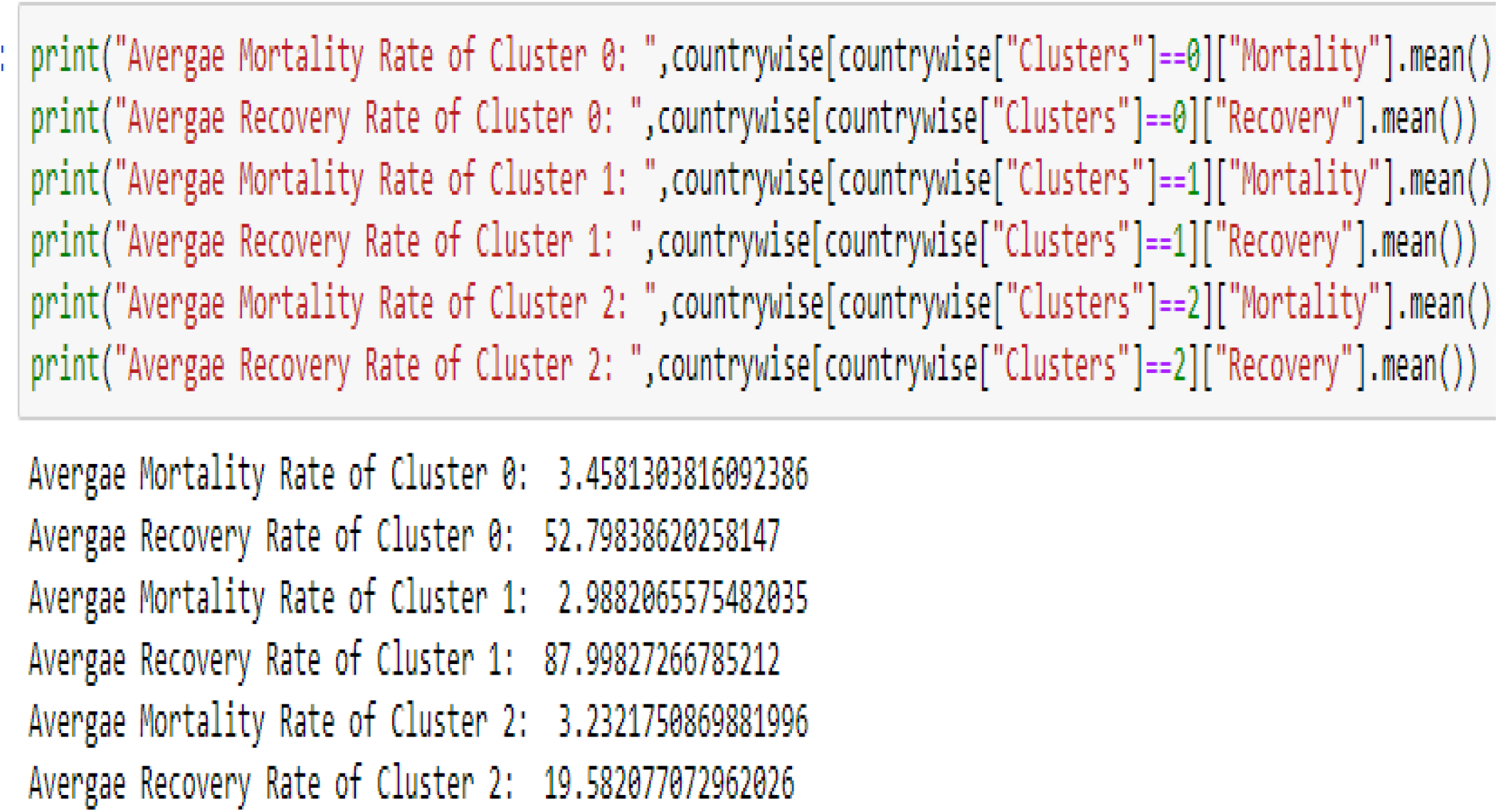

We can see countries belonging to cluster 1 are at a comparatively safer zone with low mortality rate and high recovery rate. Countries belonging to cluster 1 are India, Peru, Chile, Pakistan, Bangladesh, USA and Russia.

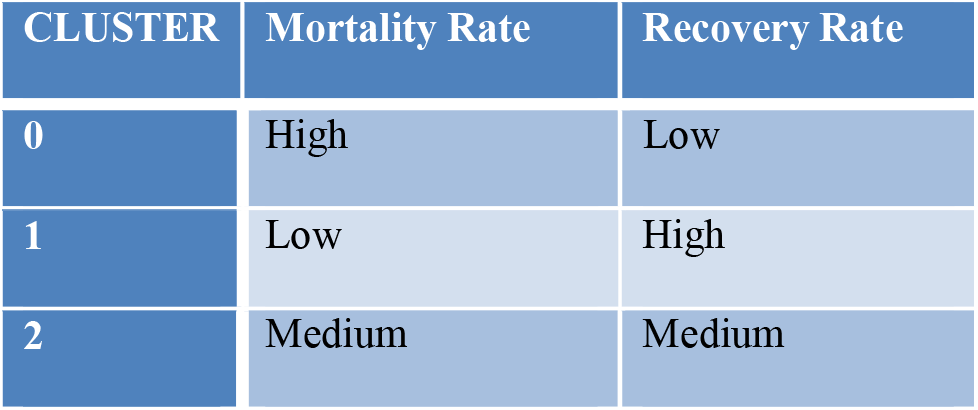

A gender distribution analysis reveals that males are more likely to be diagnosed with Covid-19

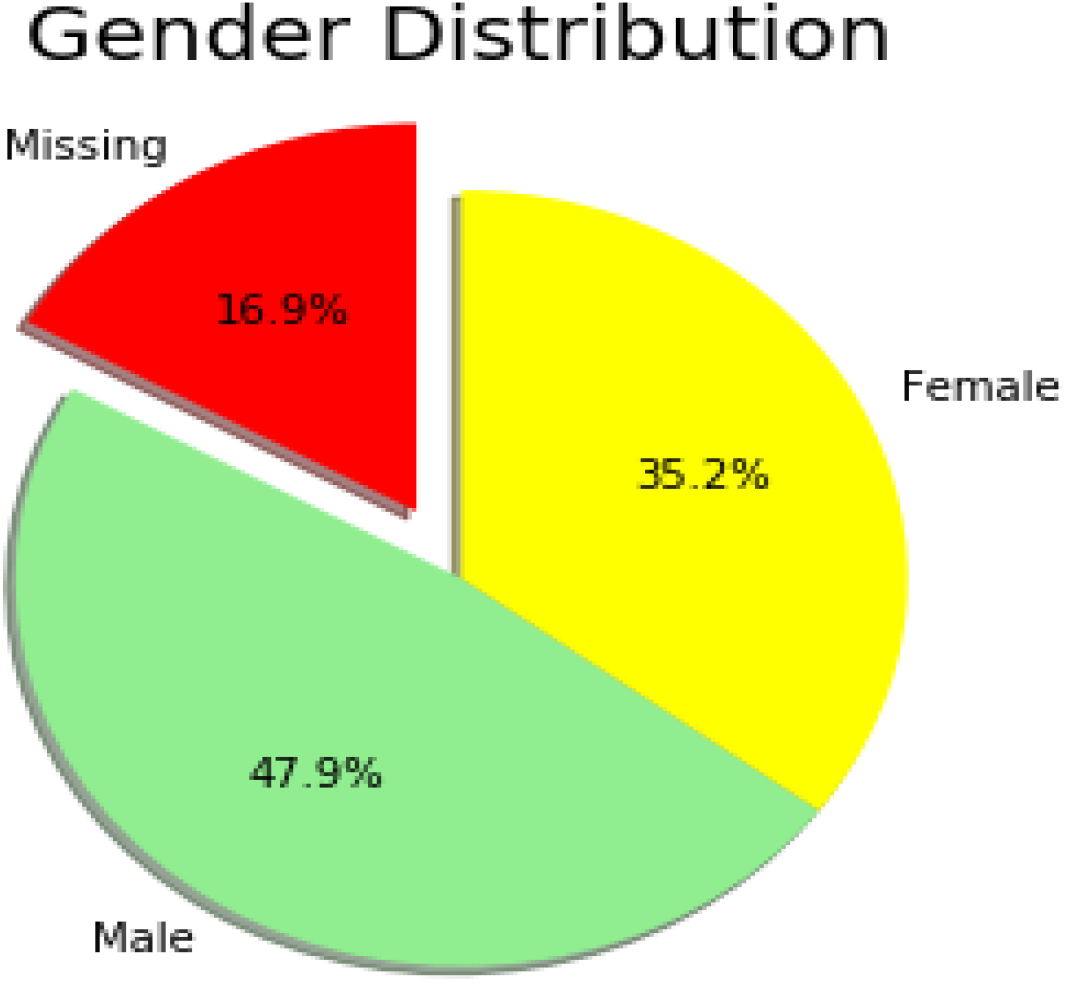

## Indian Scenario

Drilling down to the present scenario of India-data available from various sources reveals that covid-19 reached India at a little later stage. The first case was diagnosed on 30th January and a few cases were reported on February, all of them being students returning from Wuhan,China to Kerala, India.

Age wise distribution of confirmed cases till 30th July 2020 -

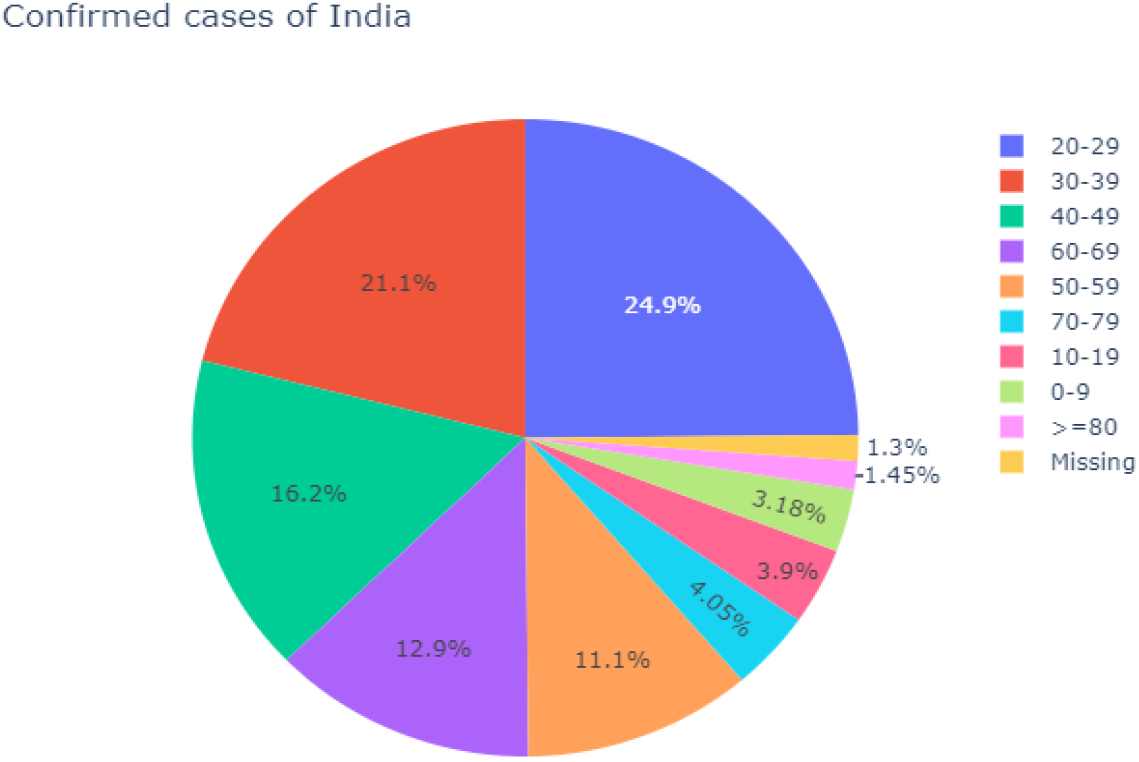

Calculating the average mortality rate and recovery rate-

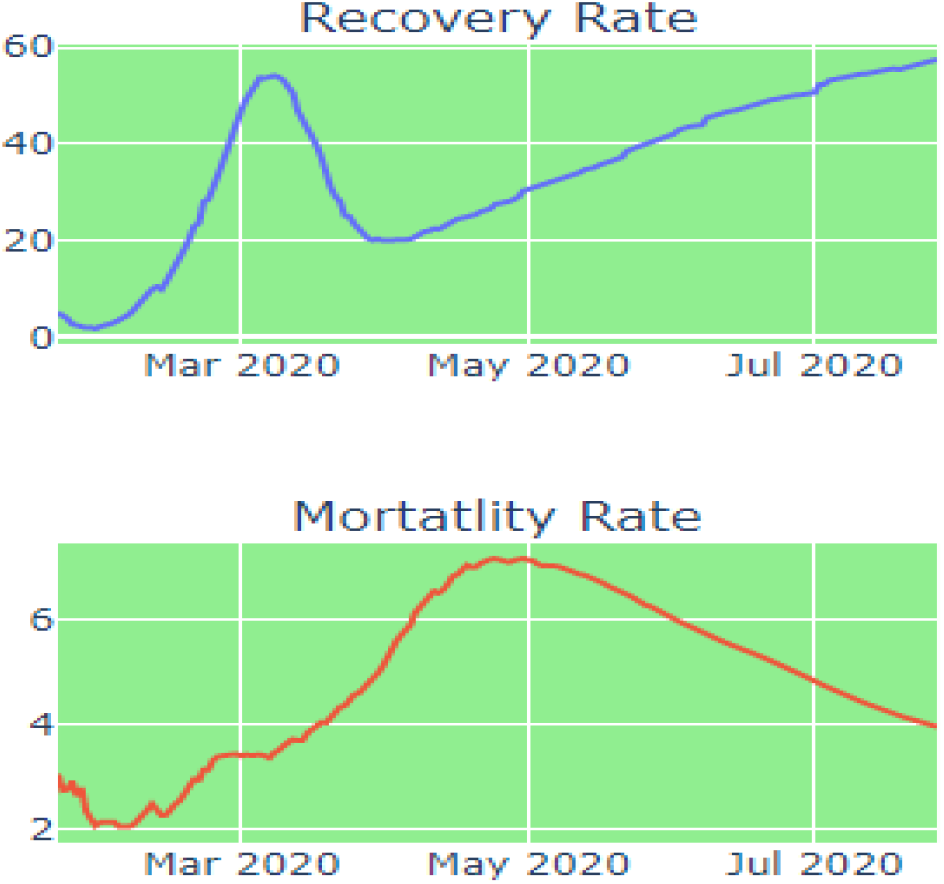

Growth factor (Graph in log scale) -

It is calculated by the number of cases confirmed yesterday divided by that of today. Using the shift function of python, we can find out a continuous trend of cases increasing with respect to the previous day. A growth factor of 1 indicates cases of yesterday and today was same while growth factor of 2 indicates doubling of new confirmed cases.

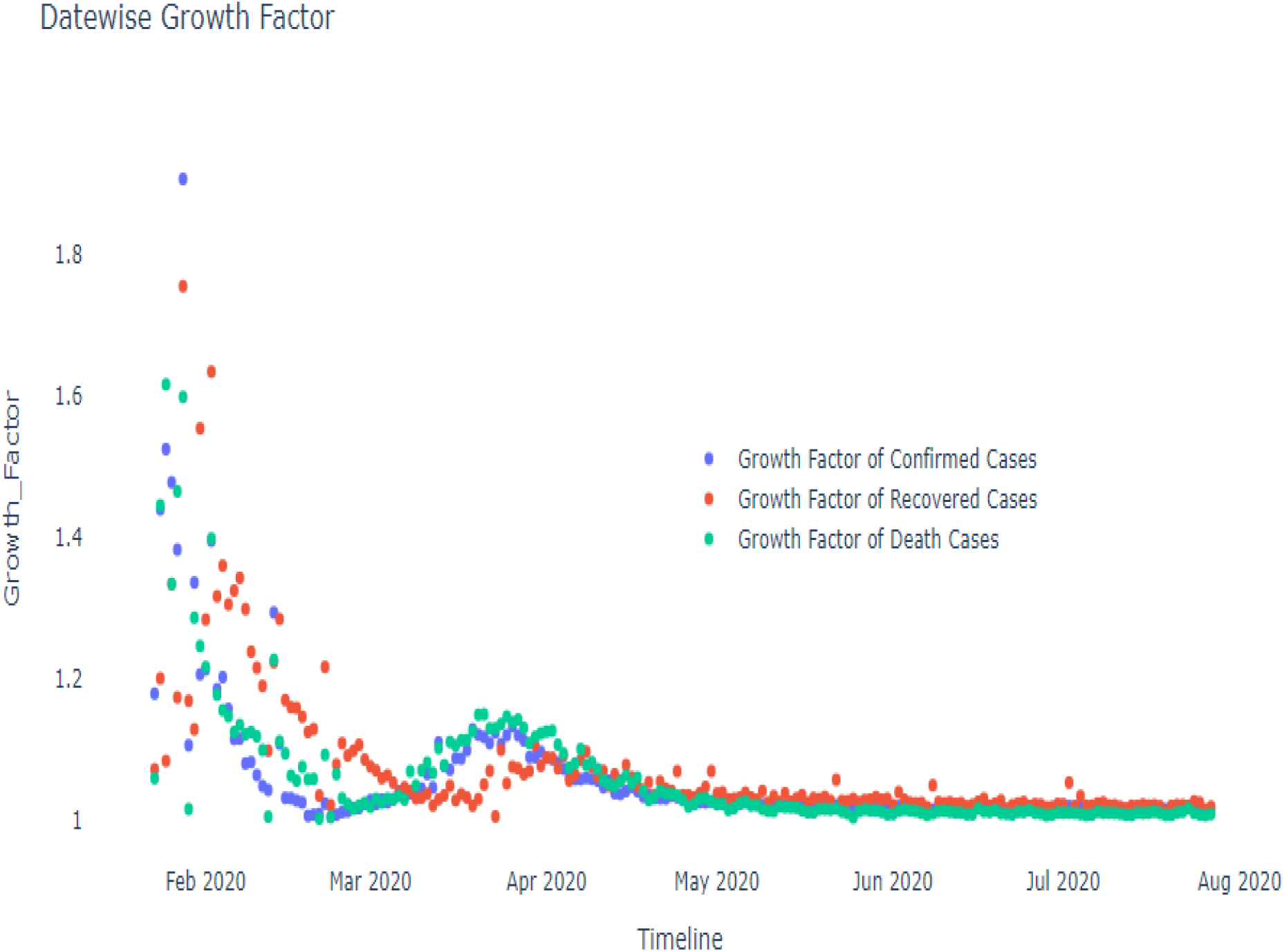

Treemap visualization of cases according to states. As of July 30th, 2020, Maharashtra, Delhi, Tamil Nadu, Gujarat where were at the top of the list.

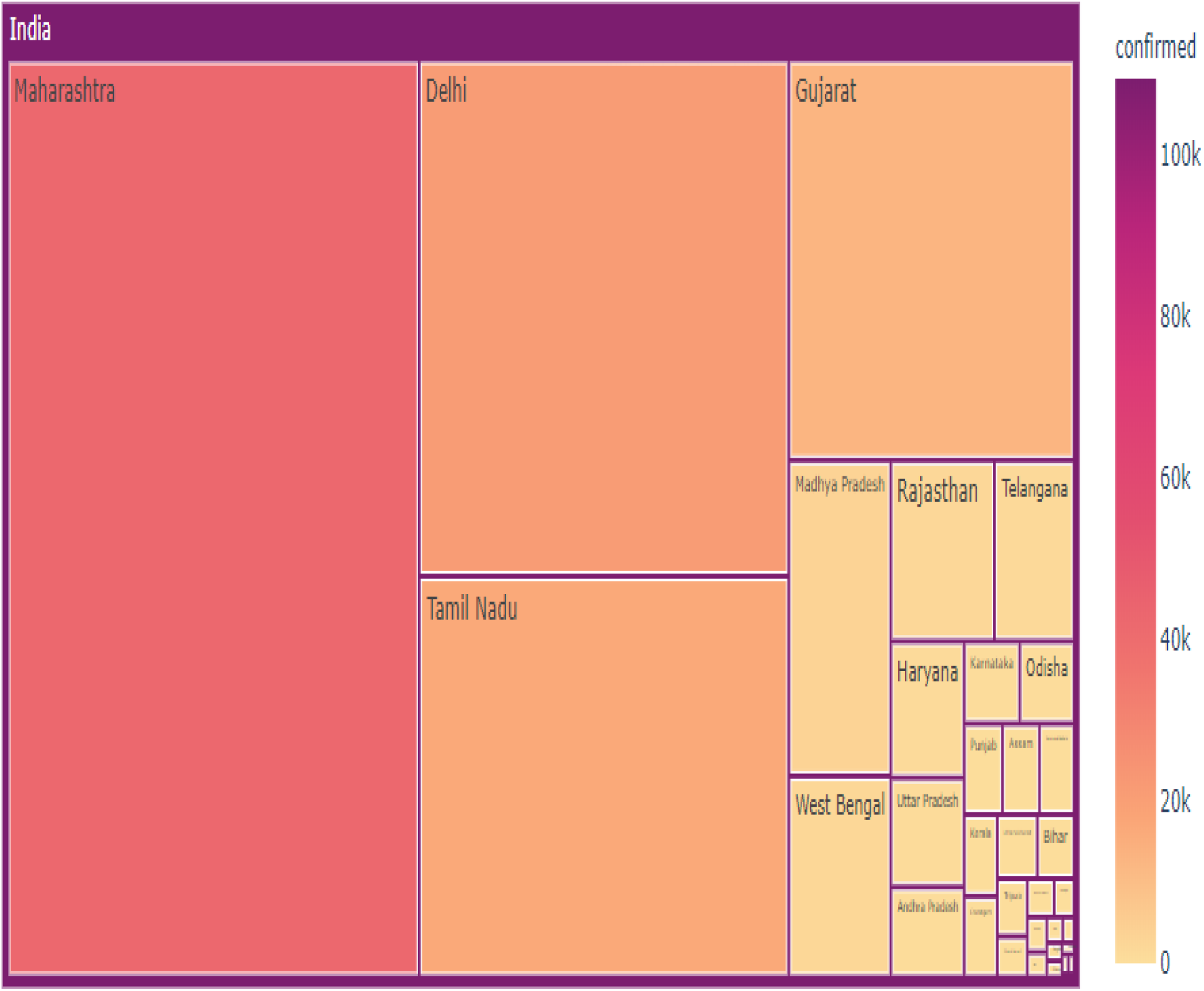

## Forecasting

^[7]^Arima (Auto-regressive integrated moving average) is a popularly used time-series prediction model. It assumes that there is a linear relationship between the variables and time. This model is very dynamic and can be run again and again over a course of time period.

^[8]^Prophet is another forecasting model given by Facebook, which is designed to manage business problems. It has two techniques of growth forecasting-here we are using the saturated growth model. Model is very flexible to handle periodic effects.

^[9]^Linear regression model trains itself on some historic data of independent and dependent variable and predicts the future considering a linear relationship between both. Polynomial Regression uses a similar approach; only difference is the dependent variable is modeled as nth degree of the polynomial in x.

^[10]^Support vector regression is a supervised machine learning model which draws a hyper plane between the data points and creates a boundary of possible data points (high and low) in future. Support Vector Regression traditionally has huge forecasting ability.

The below figure shows that polynomial regression is a good fit for our data. Also subsequently we have the ARIMA model and the Facebook Prophet model which also fits well to our dataset. However it doesn’t give us an estimate of the date when we can expect a downfall in the number of cases. We had also tried with linear regression and support vector machine regression which did not give accurate results in model fitting.

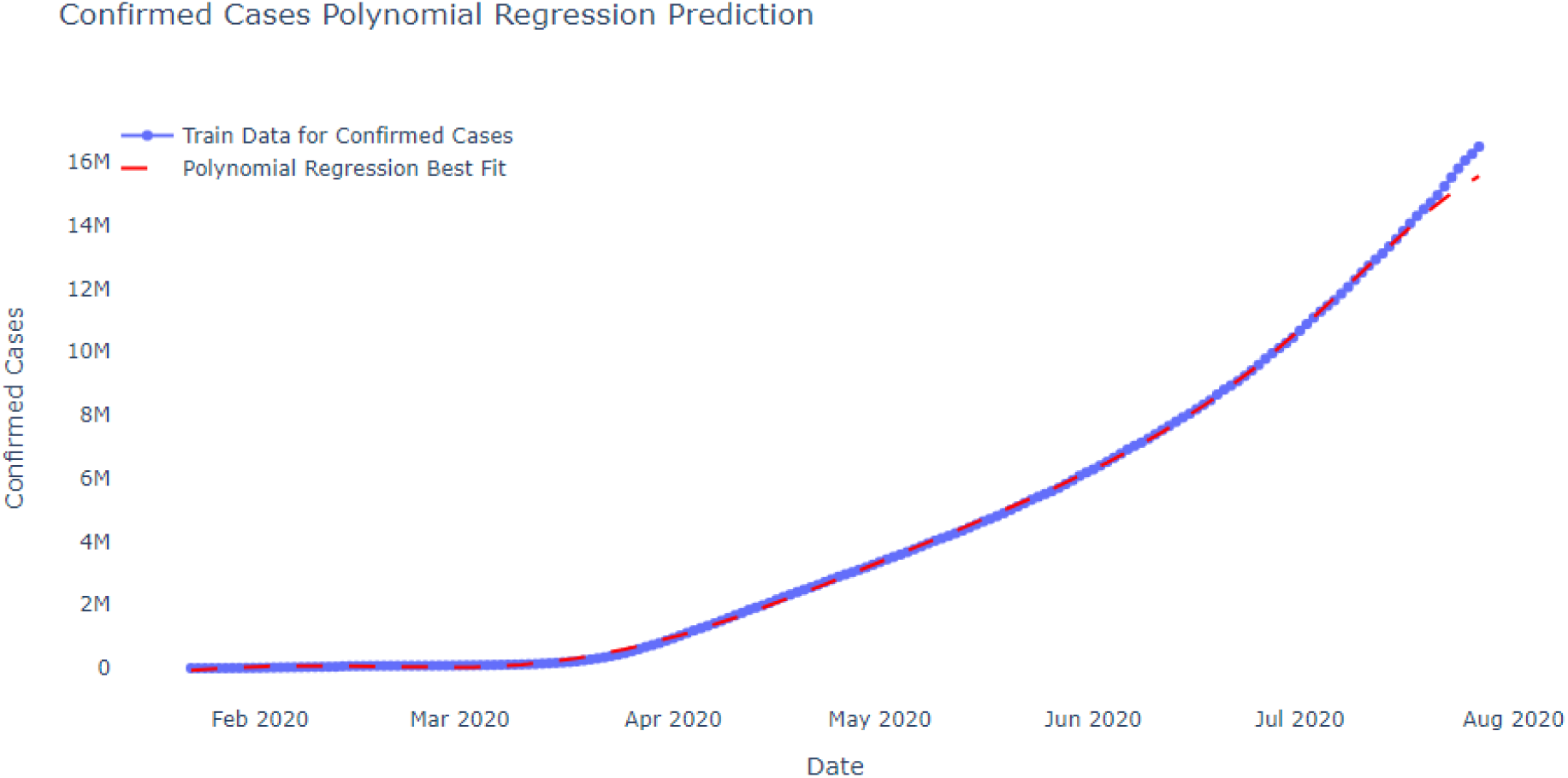

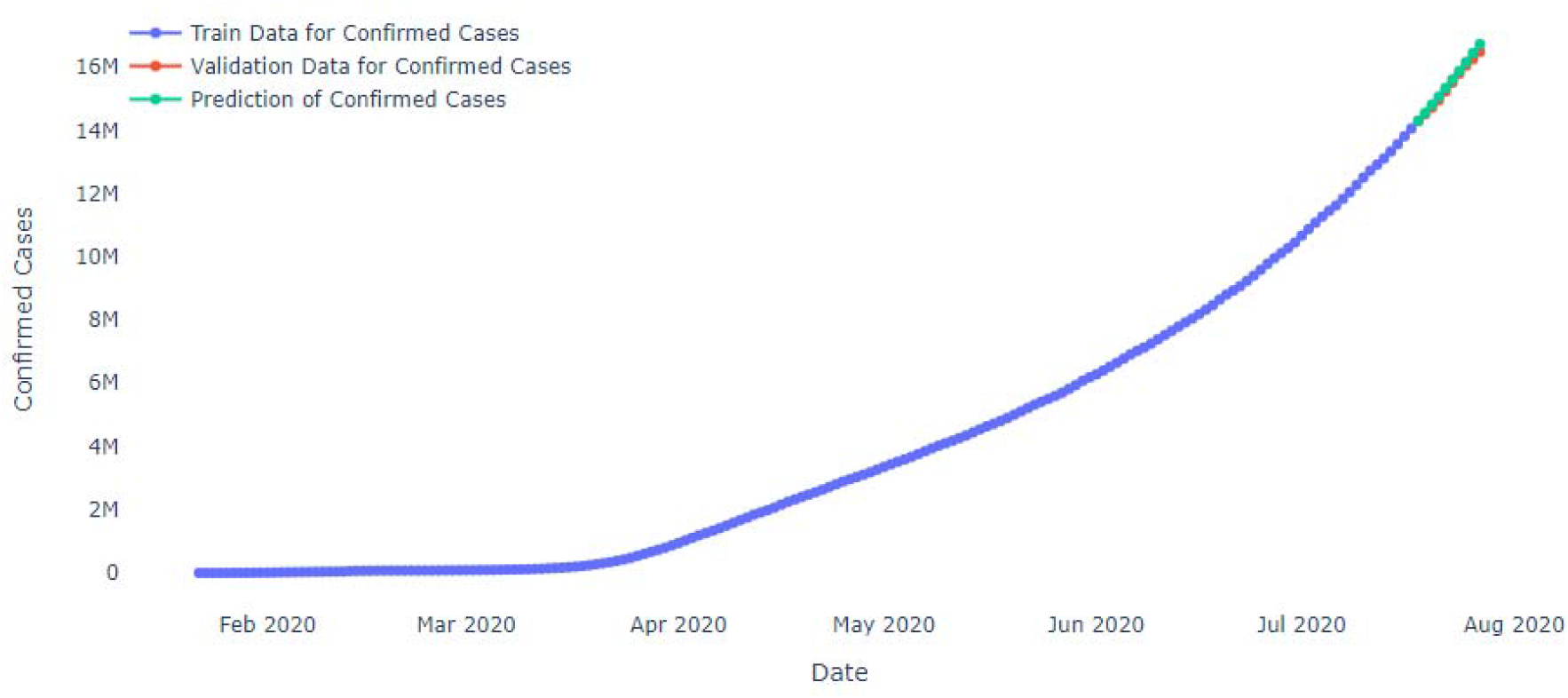

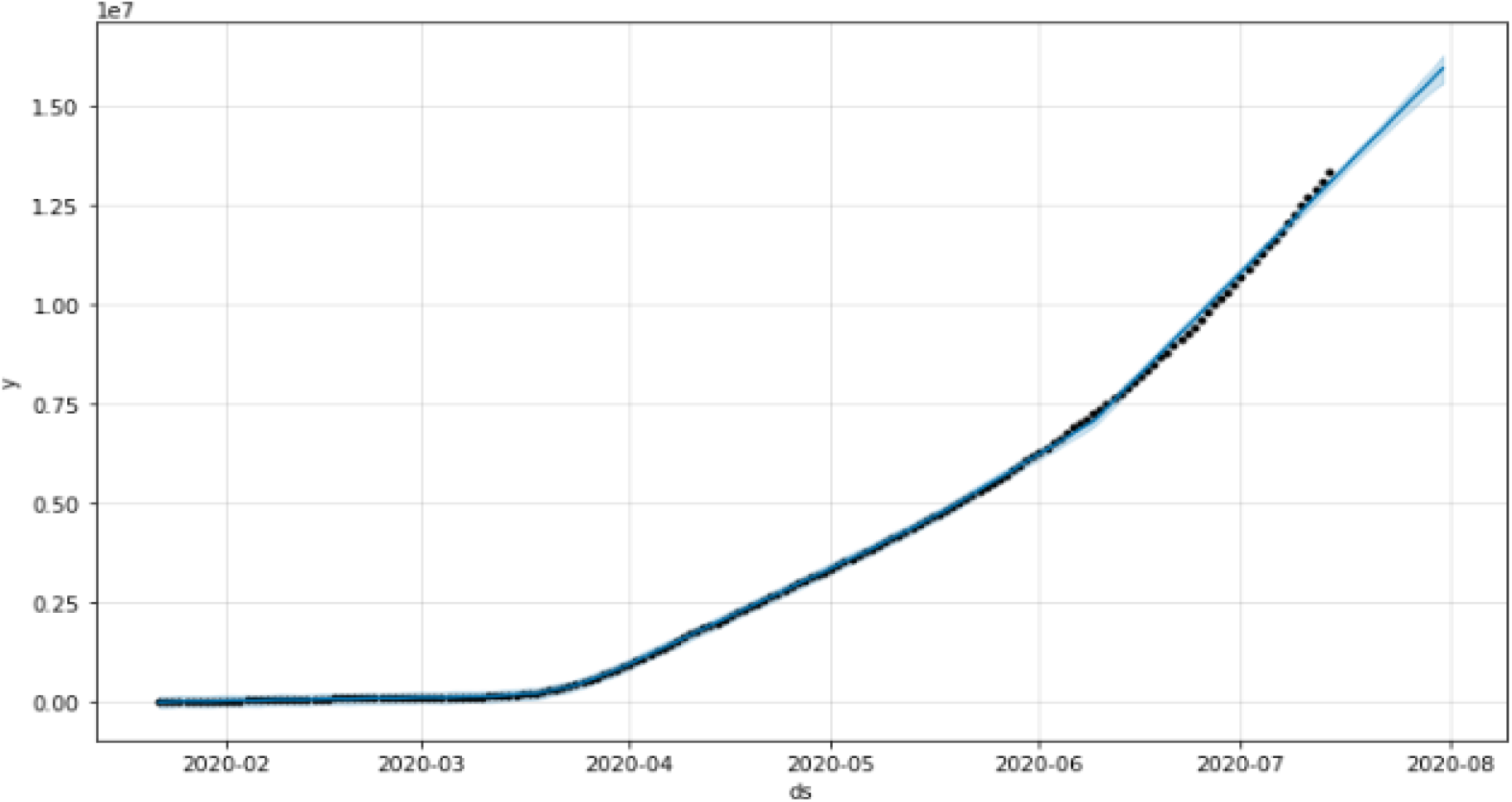

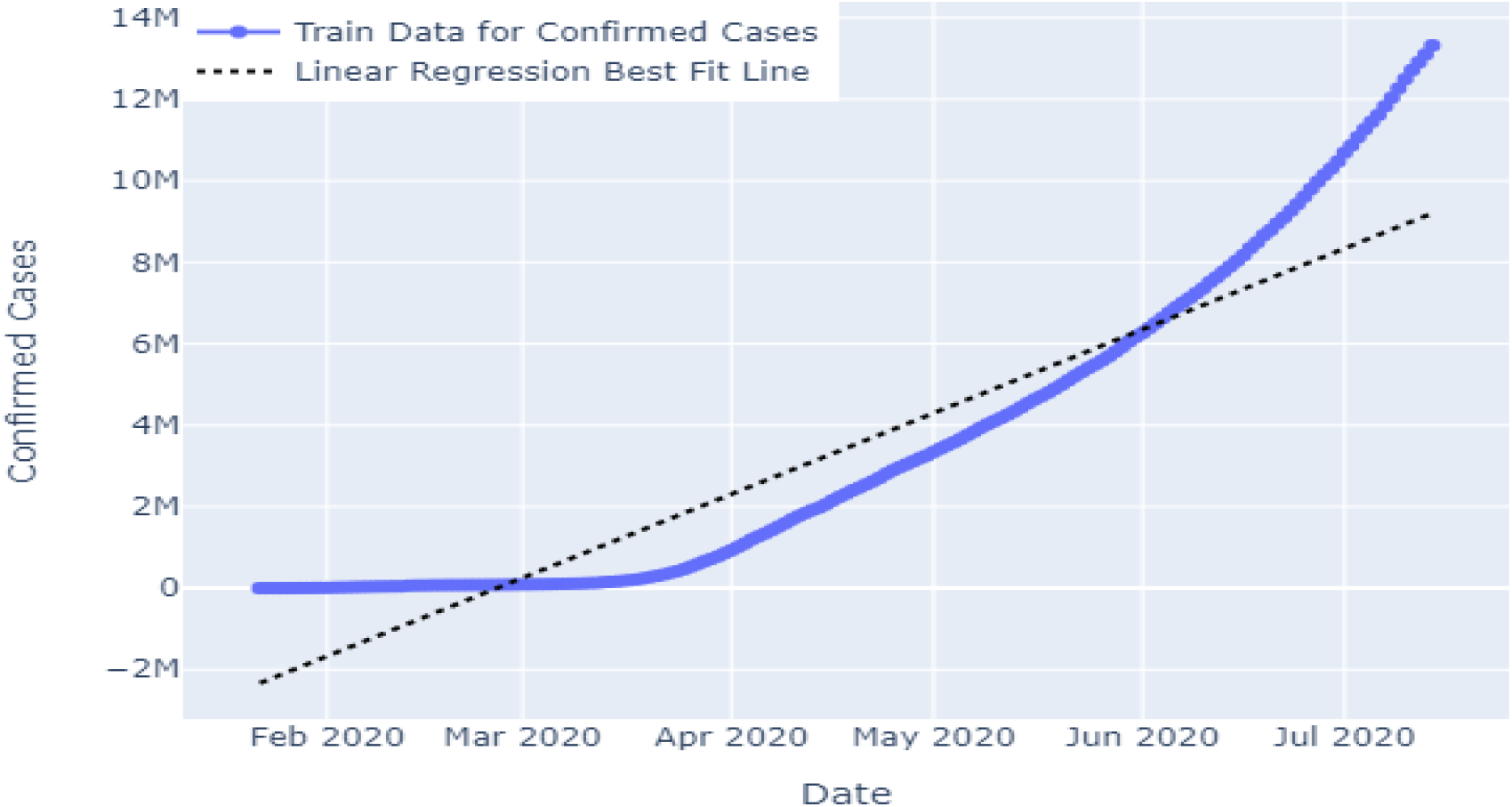

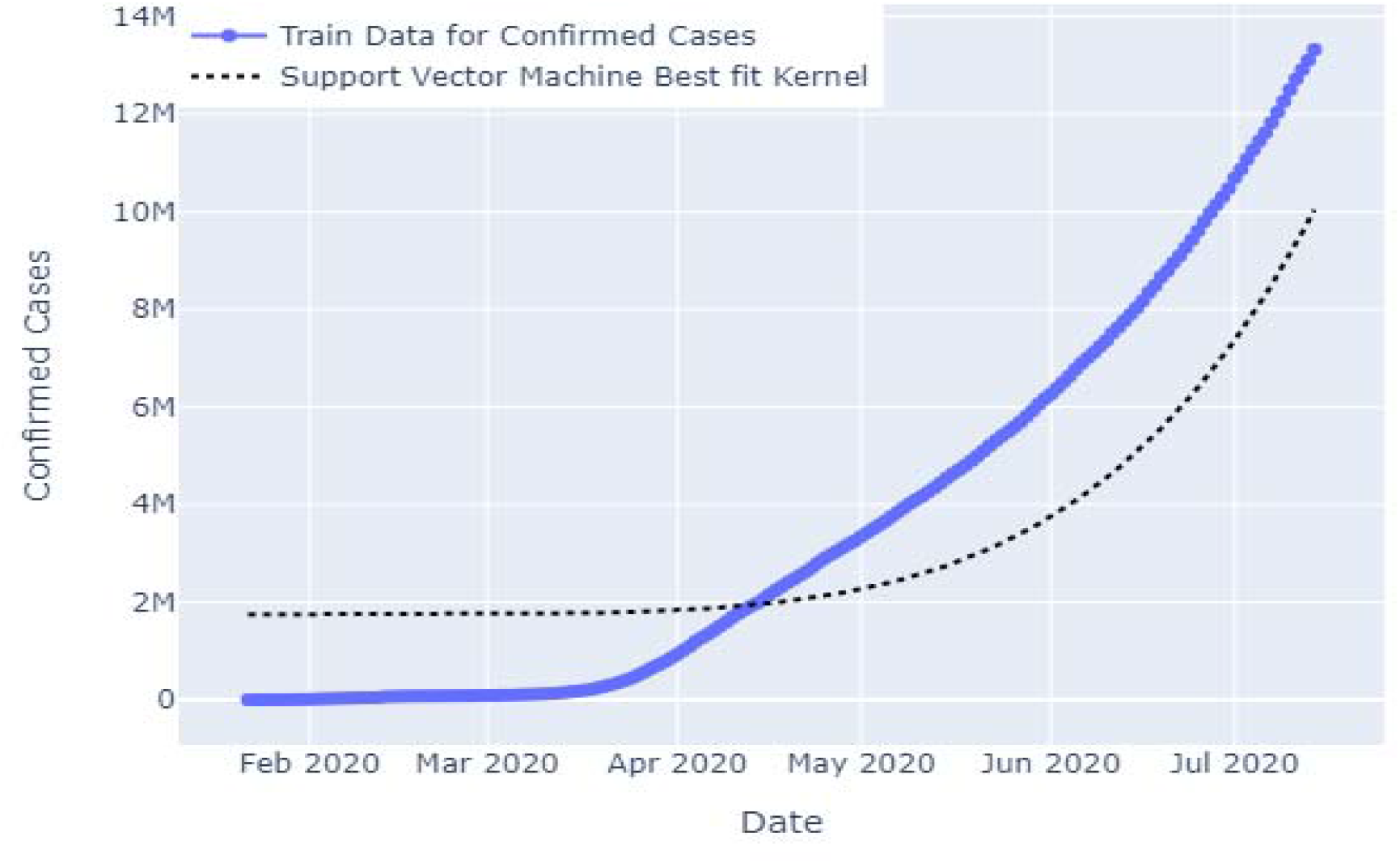

### Forecasting using LTSM (Long Short-term Memory)

^[11]^We can use LTSM to predict univariate time series problems. Usually the problems consist of a single series of observations.It is an artificical recurrent neural network architechture that takes historic data, trains the model and predicts the near future.

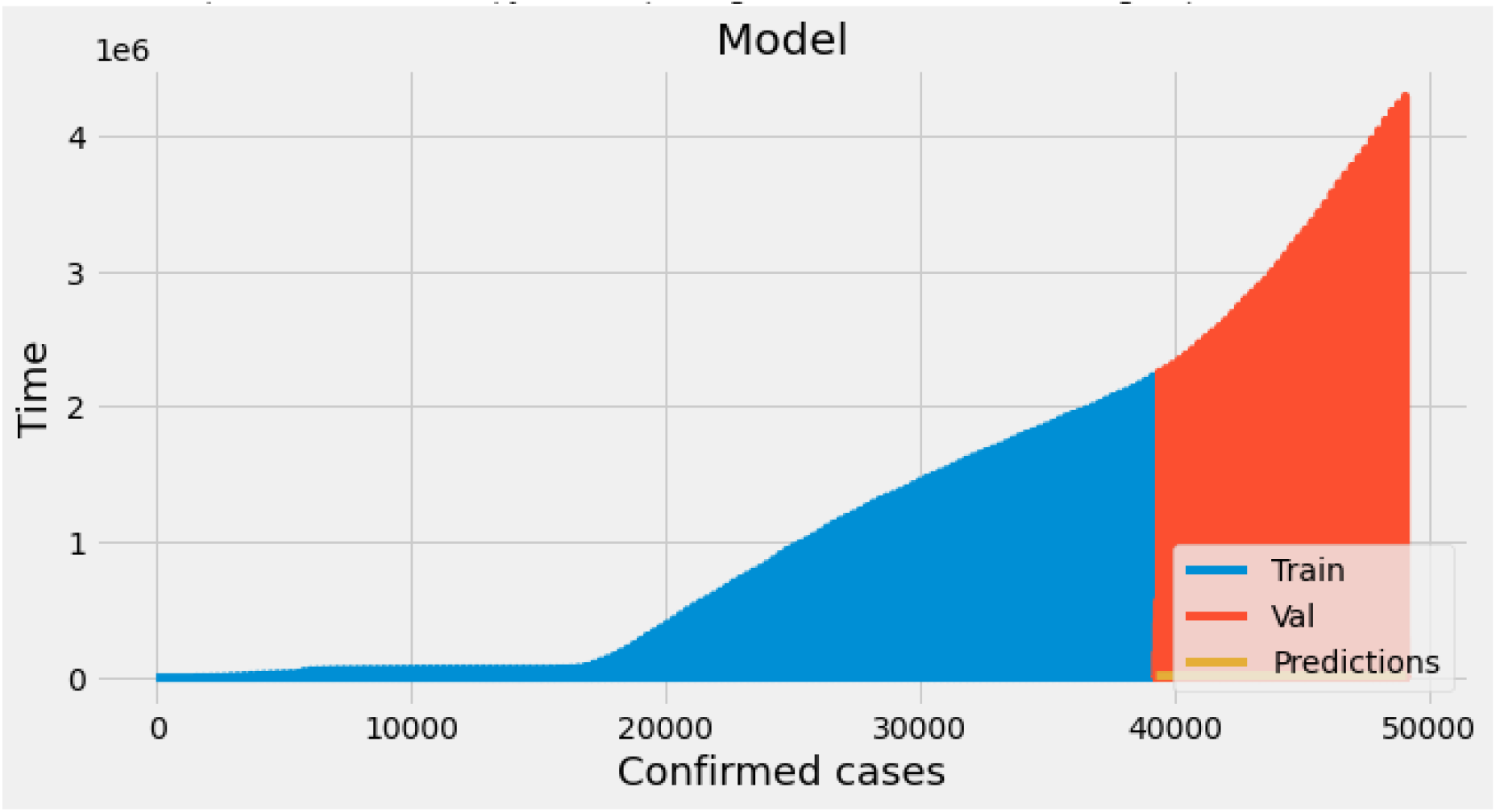

Classical methods like Arima Model, Facebook Prophet forecasting model and polynomial regression outperform machine learning methods for one step forecasting on our dataset. These machine learning and deep learning models do not deliver their promise of accurate time series forecasting and we have much more to

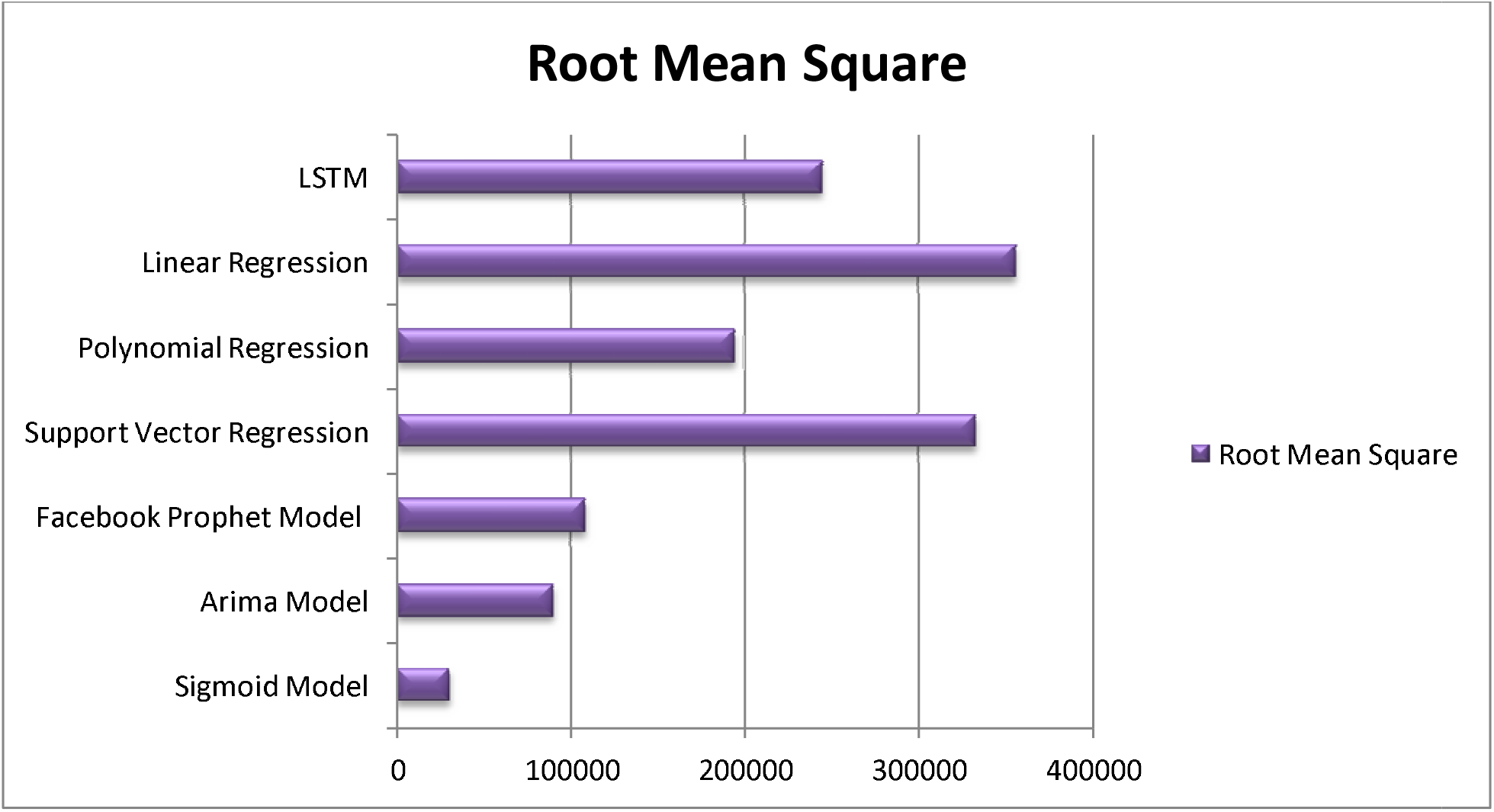

This bar chart gives us a comparative analysis of the fitness of each model. Lower the root mean square better is the model fit. The Arima Model and Facebook Prophet Forecasting Model are showing a good fit while Linear Regression and Support Vector Regression are the worst fit. This is supported by the graphical results of prediction shown above.

The Sigmoid Model has lowest Root mean square value, so it is the best fit.

<u>Forecasting using sigmoid model –</u>

The Machine Learning and deep learning models used above are traditionally used for time-series forecasting and give brilliant results elsewhere for example sales forecasting. However the case prediction of Covid-19 is different. We are currently at a growth phase when it comes to number of daily active cases; however considering the graphs of several other countries, in India also we are expecting a point from which the curve will fall. This kind of scenario can be best analyzed and supported by a Sigmoid model. Even China’s data also resembles sigmoidal shape.

Fitting sigmoid function in the data –

y= ———

a - sigmoidal shape progress of infection (better if small in our case)

b - The point where sigmoid star to flatten from steepening - midpoint of segment when rate of increment of cases will slow down.

c - The maximum value (maximum number of infected patients)

## Results

<u>Python calculation and overview of the data -</u>

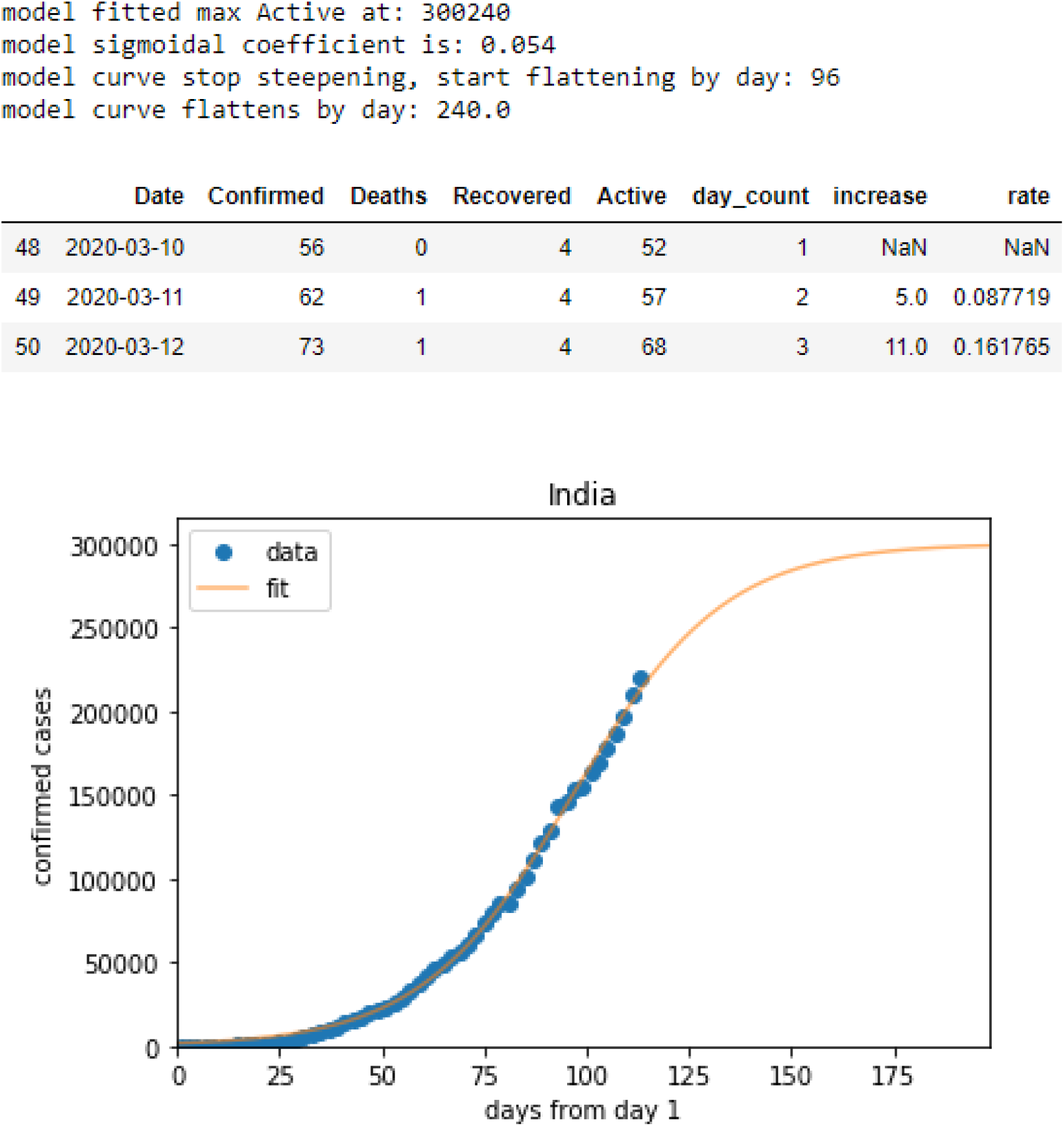

The model predicts maximum active cases at 300240.

The curve flattens by day 240 i.e. 19^th^ September and after that the curve goes down and the number of active cases eventually will decrease.

## Conclusion -

There are a lot of research works going on with respect to vaccines, economic dealings, precautions and reduction of Covid-19 cases. However currently we are at a mid-Covid situation. India along with many other countries are still witnessing upsurge in the number of cases at alarming rates on a daily basis. We have not yet reached the peak. Therefore cuff learning and downward growth are also yet to happen. Each day comes out with fresh information and large amount of data. Also there are many other predictive models using machine learning and deep learning that have been used to forecast. However the Sigmoid model only gives us an accurate representation of date when we can expect the peak. At the end of the day it is only the precautionary measures we as responsible citizens can take that will help to flatten the curve. We can all join hands together and maintain all rules and regulations strictly. Maintaining social distancing, taking the lockdown seriously is the only key. This study is based on real time data and will be useful for certain key stakeholders like government officials, healthcare workers in preparing a combat plan along with stringent measures. Also the study will help mathematicians and statisticians to predict outbreak numbers more accurately.

## Data Availability

All the data is I used are available online. Also I have mentioned the details in the references.

## Highlights of the paper - Results of Foecasting Models

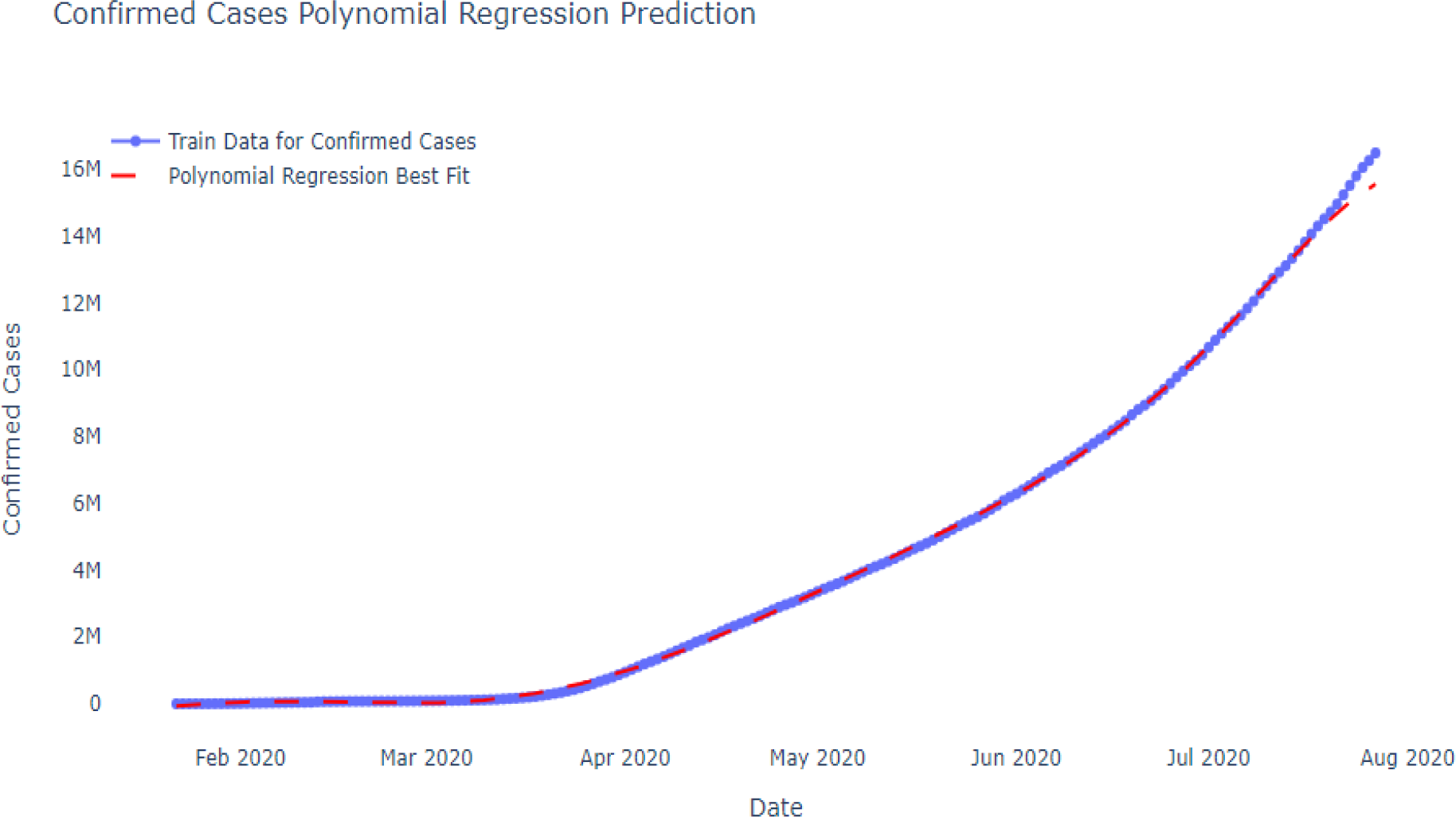

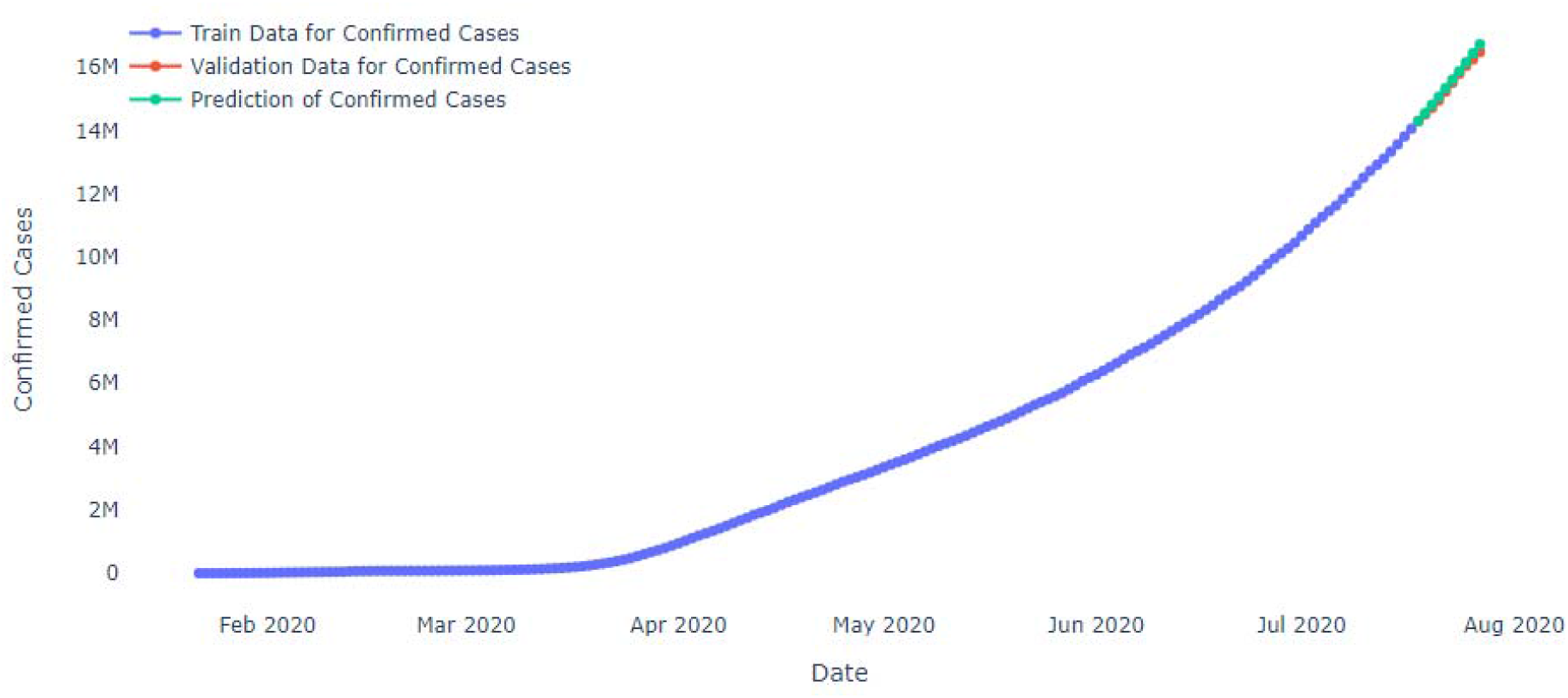

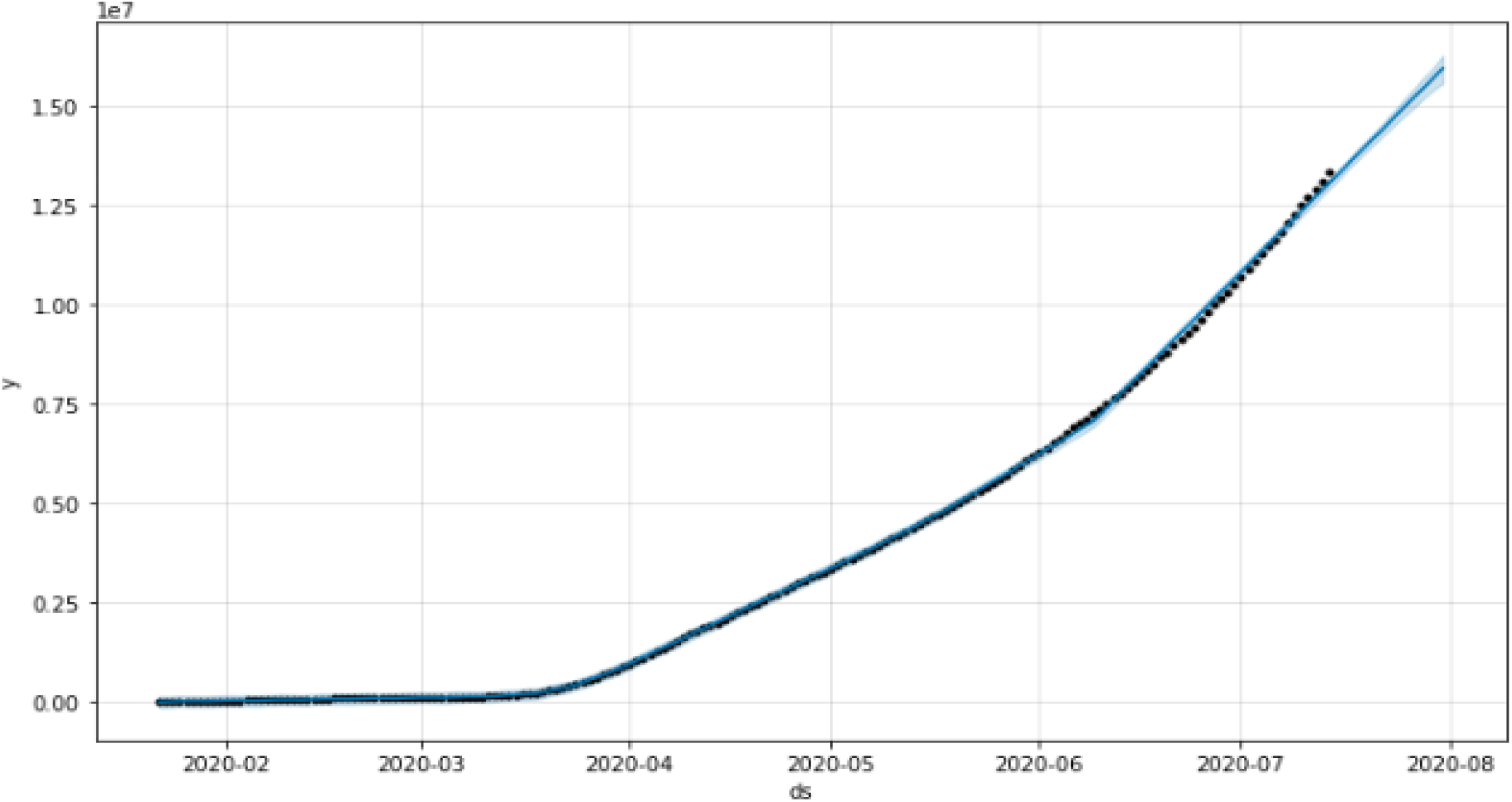

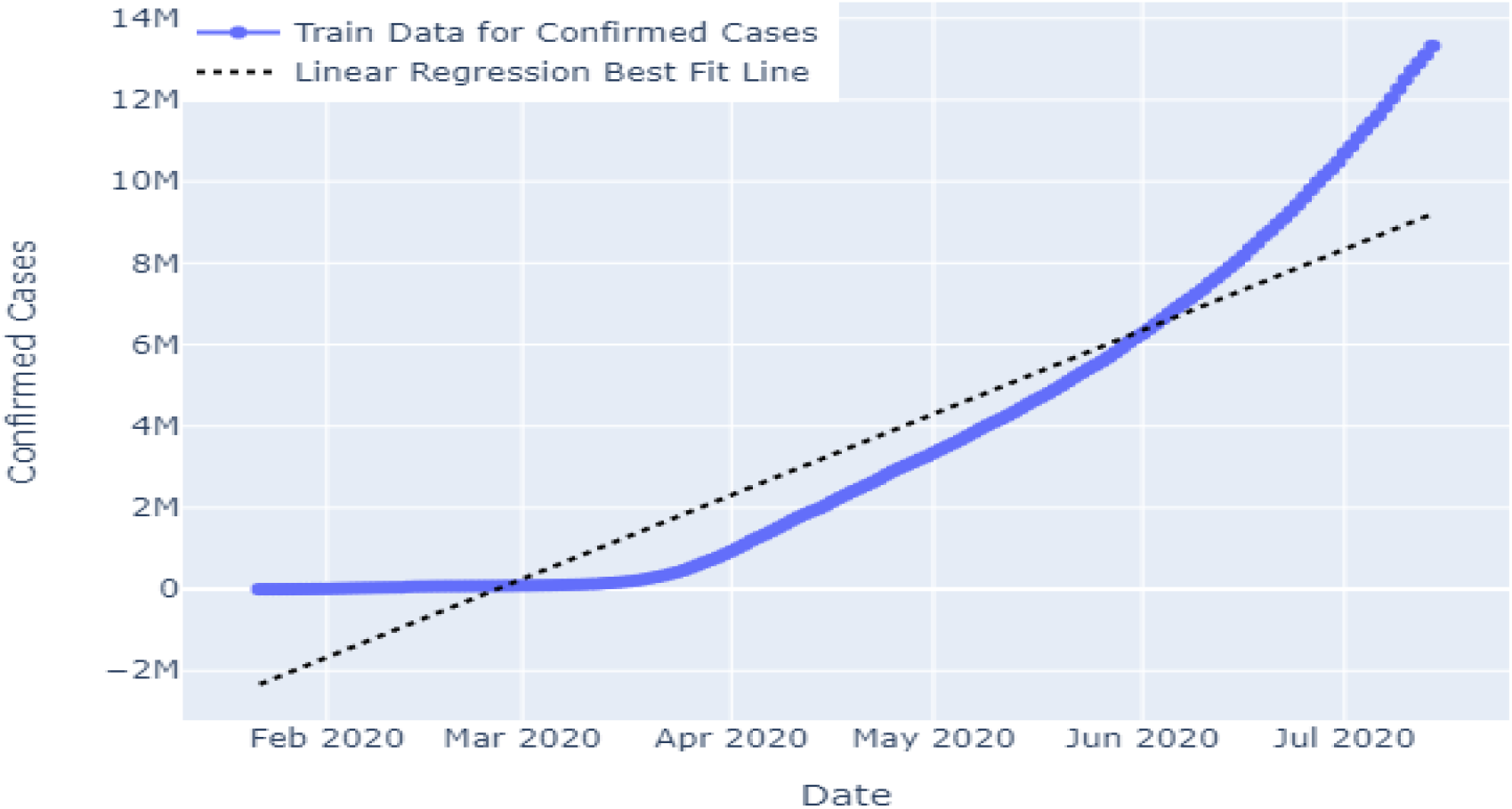

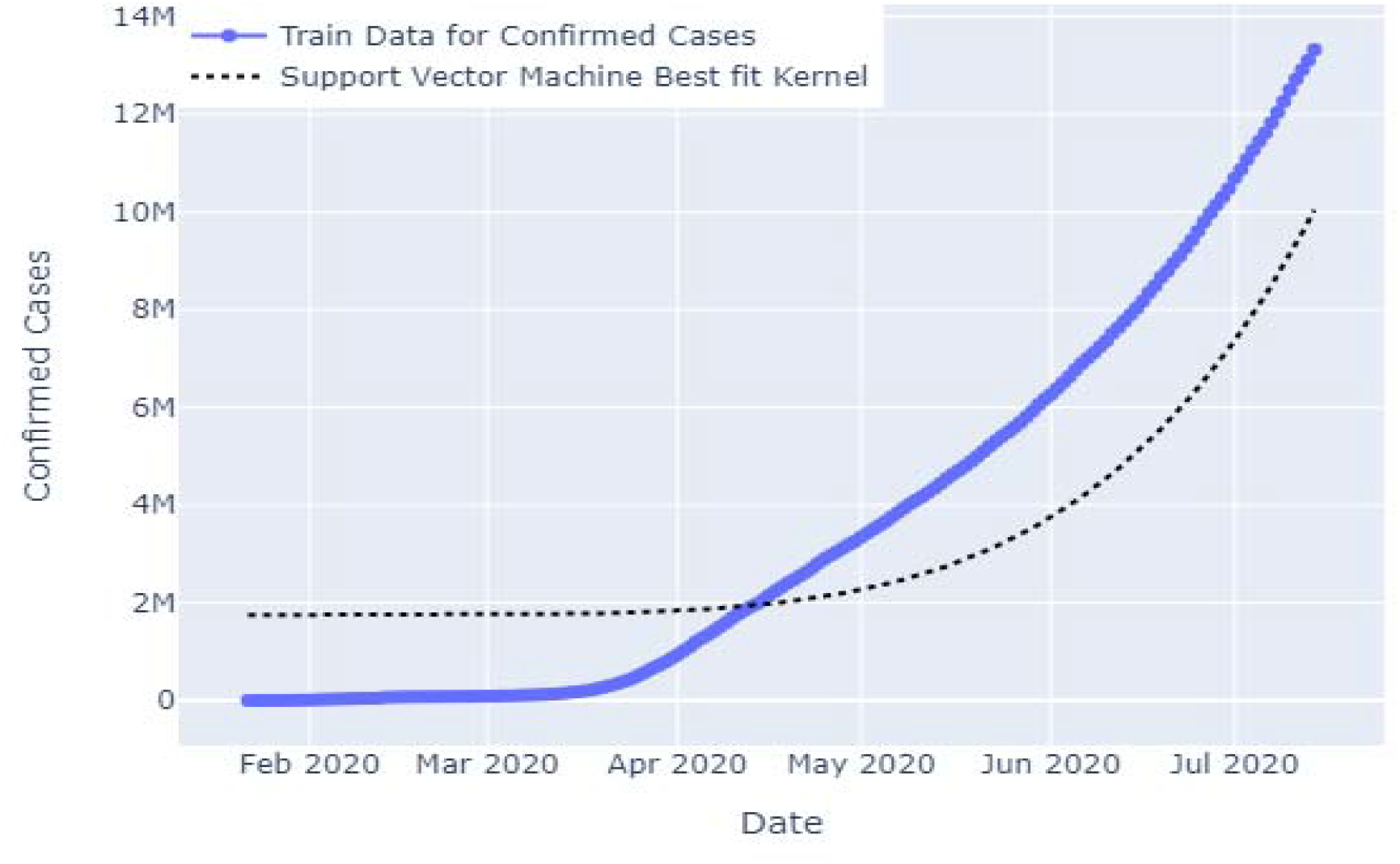

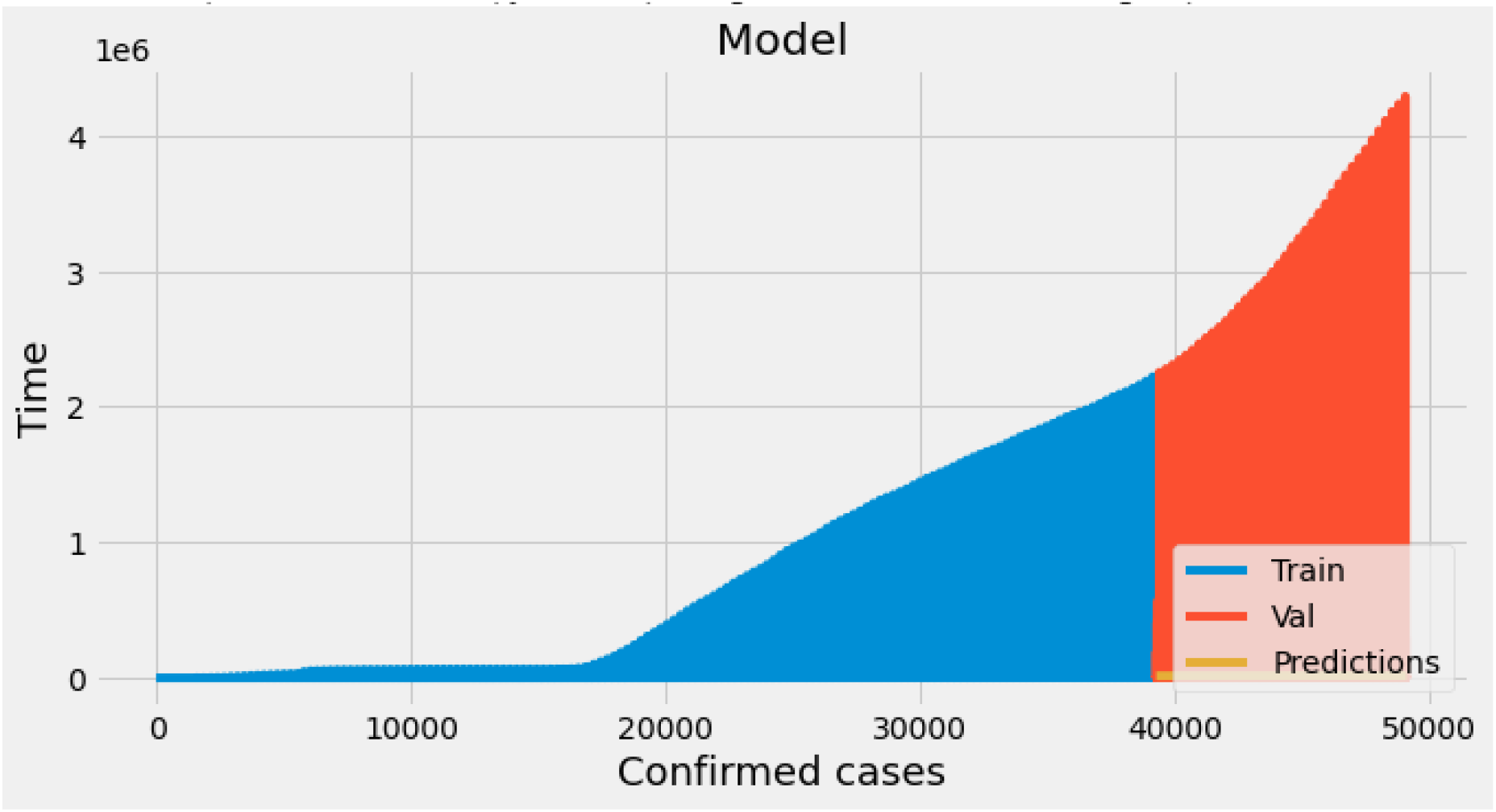

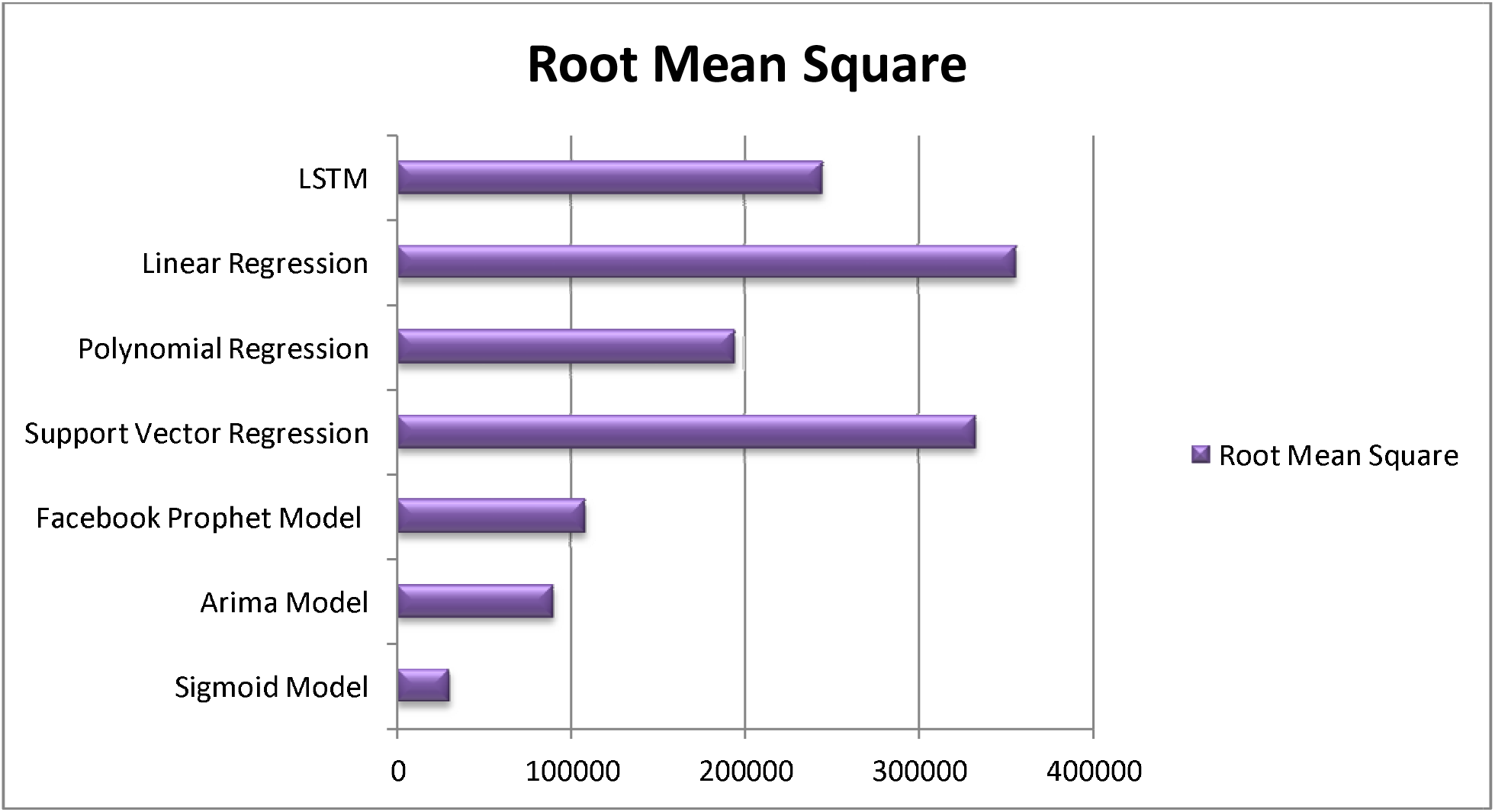

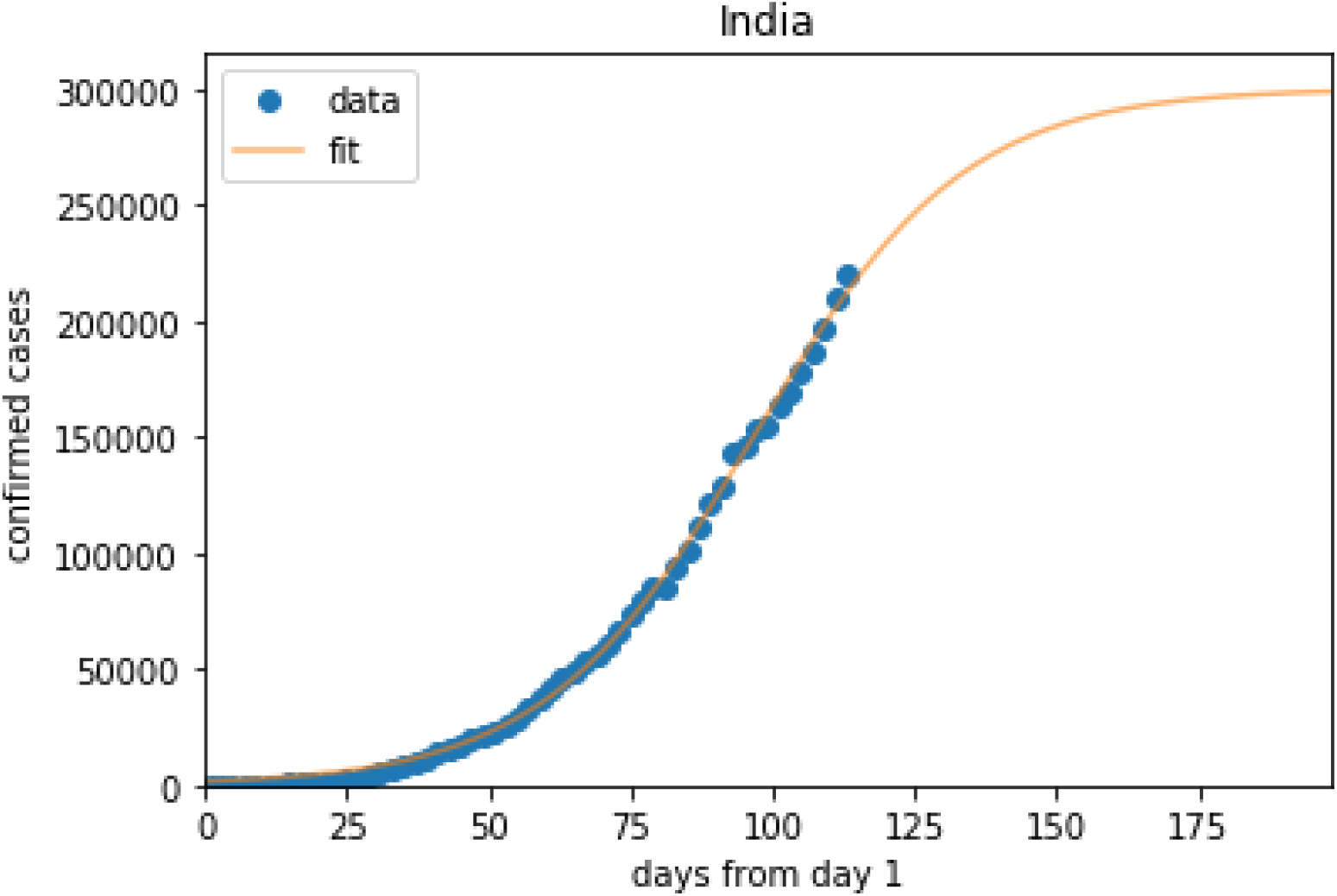

